# Reconstruction of historical malaria transmission in Senegal using multiplex sero-catalytic models

**DOI:** 10.1101/2025.11.17.25340391

**Authors:** Gaëlle Baudemont, Thomas Obadia, Laura Garcia, Camille Lambert, Françoise Donnadieu, Fatoumata Diene Sarr, Joseph Faye, Cheikh Sokhna, Inès Vigan-Womas, Aissatou Toure-Balde, Chris Drakeley, Makhtar Niang, Michael White

## Abstract

The advent of multiplexing technologies, allowing antibodies to hundreds of antigens to be measured in a single test, has led to enormous increases in the amount of data generated by serological surveys. New modelling methods are required to exploit this data. This study extends serocatalytic models to consider up to three antibody responses targeting the same pathogen simultaneously. These models were fitted to data from cross sectional serological surveys of *Plasmodium falciparum* malaria in the Senegalese villages of Dielmo and Ndiop, and model predictions were validated against 22 years of longitudinal epidemiological data. The most accurate reconstruction of historical clinical incidence of *P. falciparum* was provided by a combination of antibodies to Apical Membrane Antigen 1 (AMA1) and Glutamate-Rich Protein (GLURP). This model estimated a 76% (95% CrI: 61% - 86%) drop in transmission in 2004 (95% CrI: 2001 – 2008) coinciding with changing anti-malarial treatment. Multiplex serocatalytic models provided more accurate estimates of past clinical incidence than singleplex serocatalytic models, with the most accurate multiplex model (AMA1 + GLURP) outperforming all singleplex models (Kruskal Wallis p < 0.01). Finally, models with three antigens did not provide more accurate estimation than models with two antigens.

**Significance Statement:** Multiplex serological assays are accelerating the uptake of serology as a surveillance tool for malaria. However, there has been a lag in the development of analytic tools for the analysis of this rich multiplex data on measurements of antibodies to multiple malaria antigens. In this study, new mathematical models are developed to account for multiplex data. We demonstrate that these models provide more accurate estimates of past malaria transmission in Senegal, with lower levels of uncertainty compared to models based on data from antibodies to a single antigen. By providing more reliable estimation, we anticipate that our work can form the basis of an important new tool for malaria surveillance.

## Introduction

The WHO warns that the malaria burden is increasing compared to the pre-COVID pandemic period (1), with more than 600,000 deaths reported, the majority of which occur in sub-Saharan African countries (1). 90% of deaths due to malaria are caused by *Plasmodium falciparum* (2), with young children and pregnant women being most affected (3,4). Overall, 40% of the world’s population live in malaria endemic countries (5). While this result comes partly from the interruption of public health systems during the pandemic, it can also be linked to global warming, notably extreme weather events (flooding, heatwaves), the apparition of drug resistance against artemisinin-based combination therapies (6), and a more challenging environment for funding of global health interventions. This emphasizes the necessity of sustained research efforts toward malaria elimination and the development of improved surveillance tools to better inform intervention strategies and policy decisions.

Serological surveillance is an important tool to assess malaria transmission (7,8): it can be implemented in low resource areas and can be more reliable than Entomological Inoculation Rate (EIR) in low-endemic and pre-elimination settings (9,10). Serological assays detect antibodies induced by both recent and past infections (11), as well as asymptomatic infections, allowing for a detailed overview of population-level exposure. Technologies such as multiplex bead-based assays allow for simultaneous quantification of up to 500 antibody responses from a few microliters of plasma (12). When compared to traditional Enzyme Linked Immuno Assays (ELISA), multiplex serological assays implemented on Luminex platforms have a broad dynamic range and robust reproducibility (13). In the case of *P. falciparum*, many antibodies have been described from the entire life cycle of the parasite and validated for serological surveillance (7,12,14,15). Differences in sensitivity, specificity, longevity and biological relevance allow antibodies to be used for different objectives, from diagnostic development to assessment of vaccine candidates (16).

Serocatalytic models have been demonstrated to efficiently reconstruct historical exposure to infectious diseases by fitting to age-stratified seroprevalence (17,18). They are especially useful in places where clinical surveillance is sparse or asymptomatic infections prevail. Data requirements are minimal, typically just age and seropositivity status. Sample sizes are dependent on transmission intensity, but typically a few hundred samples is sufficient (19). However, the process of dichotomization of continuous antibody measurements can be challenging as there is no one-size-fits-all method for selecting a cut-off for sero-positivity (*White et al. Whose line is it anyway? Defining sero-positivity cut-offs for infectious disease surveillance - Accepted subject to minor revisions at J Inf Dis*.). Serocatalytic models account for long term trends of cumulative exposure and tend to smooth out seasonal effects. While the most basic form of the model considers a constant transmission setting, it can be modified to capture changing transmission and waning immunity. Although there have been many important methodological advances for serocatalytic models, analytic tools for interpreting multiplex data are notably lacking. This study extends serocatalytic models to multiplex data and demonstrates their potential for improved serological surveillance. Multiplex serocatalytic models were applied to two Senegalese villages, Dielmo and Ndiop, in which extensive surveillance of malaria has been conducted in the past 30 years. Information on both cross-sectional serological surveys and longitudinal recording of all cases of fever due to *P. falciparum* infection were used to fit and validate these models.

## Results

### Antibody responses to malaria

More than half of the villagers were represented both in 2016 and in 2018: 311 participants and 208 respectively in Dielmo. The parity was almost perfect with 48% and 49% of males respectively. Ages ranged from 5 to 94 years old and are representative of the Senegalese age pyramid. In Ndiop, 385 participants were included in 2016 and 316 in 2018. Females were slightly overrepresented in this cohort with only 38% of participants being males. Participants were aged from 4 months old to 82 years old with a median age of 17 [1;68] years old, and 11% being children under 5 years old. Out of the Seroped cohort, 84 randomly selected samples were included. Only adults were part of this sub sample with a median age of 53 [21, 90] years old. Among the french negative controls, 44% of participants were men. Information on participants is summarised in *Table 1*.

**Table 1.**
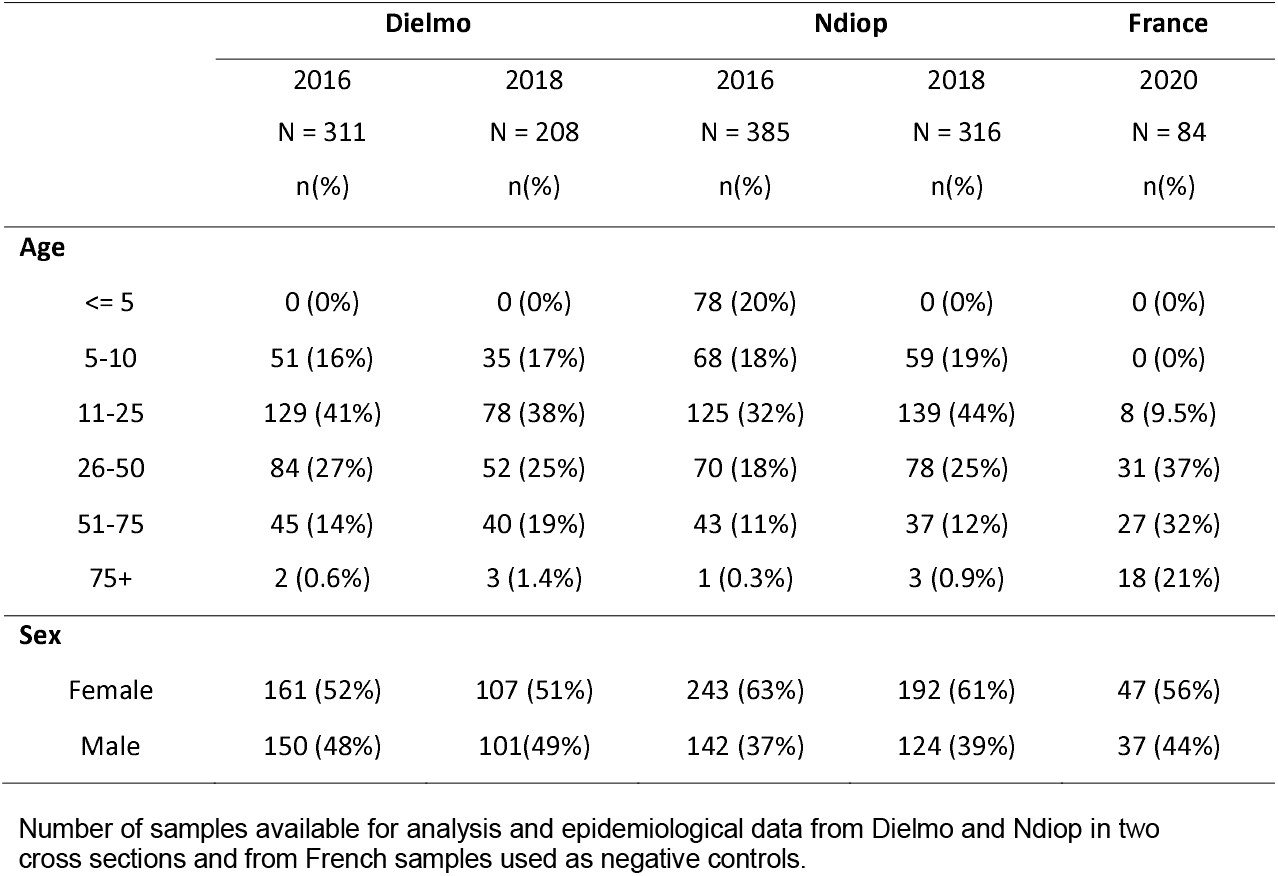
Participant characteristics.

A pattern of increased antibody response with age is observed for all tested antigens which is typical of endemic settings with continuous exposure to malaria (*Fig 1*). The distribution of the antibody responses for Dielmo and Ndiop in both 2016 and 2018 overlaps considerably. Negative control French samples had antibody responses in the lower range of the distribution. The lowest antibody responses were observed in children under five years of age from Ndiop.

**Fig 1.**
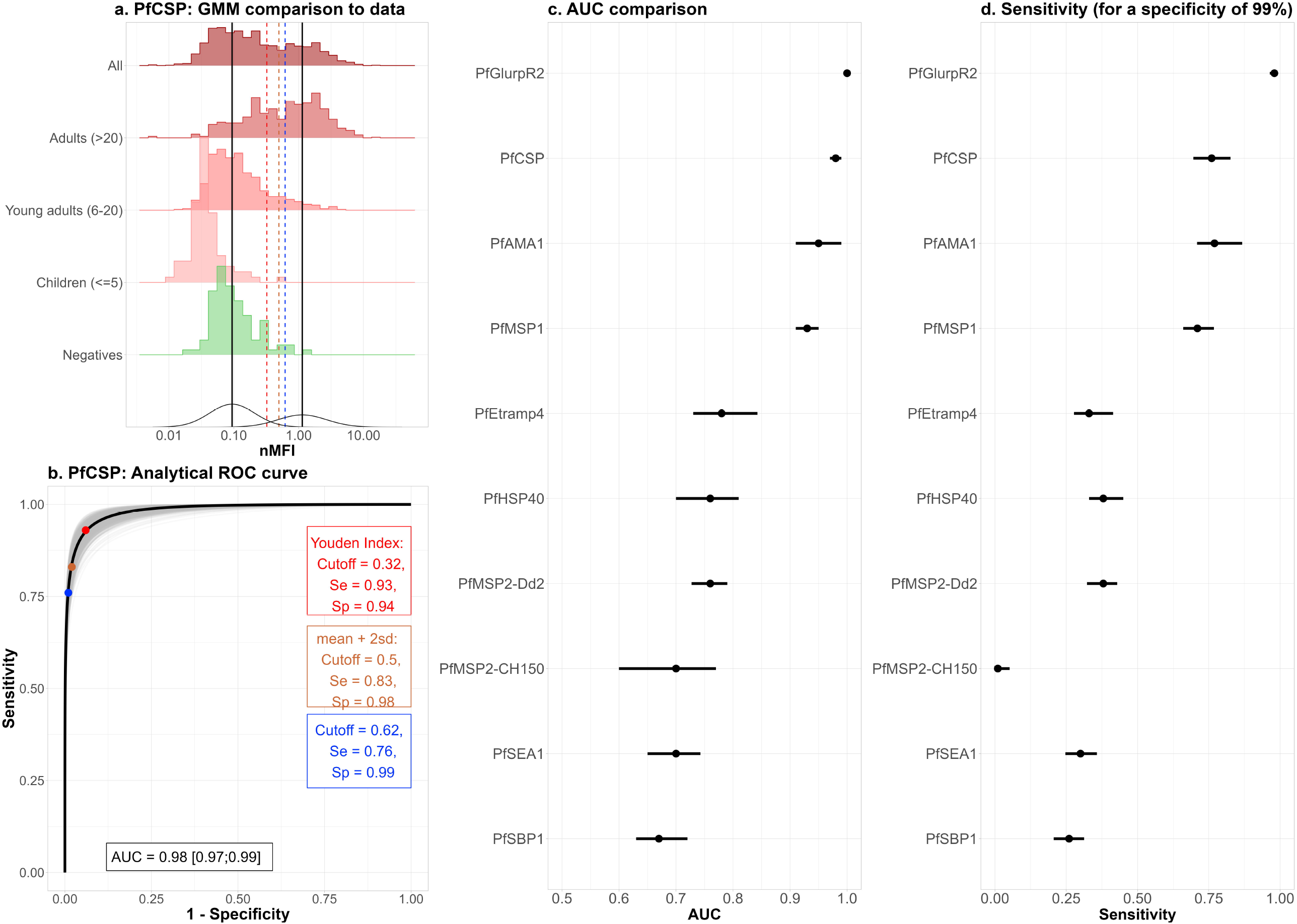
Antibody responses. Age stratified quantification using multiplex technology of ten antigens of interest, observed in the two villages and from two cross sections. Results are given in normalised Median Fluorescence Intensity (nMFI) and are compared to French negative controls. The dashed black line represents the selected cutoff to distinguish sero-positive from sero-negative participants.

### Sero-positivity classification

Gaussian Mixture Models (GMMs) applied to data from Senegalese and French samples were capable of recapturing the observed distribution of antibody responses as a combination of two components that are shown in Figure 2B for *Pf*CSP (Circumsporozoite protein), and in the supplementary information for the other antigens. The French negative control samples fall in the lower component, while for the Senegalese samples the proportion of samples falling in the upper component increases with age. For three antibody responses, *Pf*MSP-2.CH150 (Merozoite Surface Protein 2), Sporozoite Binding Protein 1 (SBP1) and Sporozoite Exported Antigen 1 (SEA1), the degree of overlap of the positive and negative distributions is too great to allow for informative classification of the samples (AUCs ≤ 0.7 *Figure* 2.C and Supplementary S1-S3) and *Supplementary S1-S3*). These antigens are not well suited to discriminate between participants that have been exposed or not to *P. falciparum*. Four antigens *Pf*GLURP.R2 (Glutamate Rich Protein C-terminal repetitive segment), *Pf*CSP, *Pf*AMA1 (Apical Membrane Antigen 1), and *Pf*MSP1 (Merozoite Surface Protein 1), have bimodal distributions and are able to classify samples (AUCs ≥ 0.9 *Fig 2* and *Supplementary S7-S9*).

**Fig 2.**
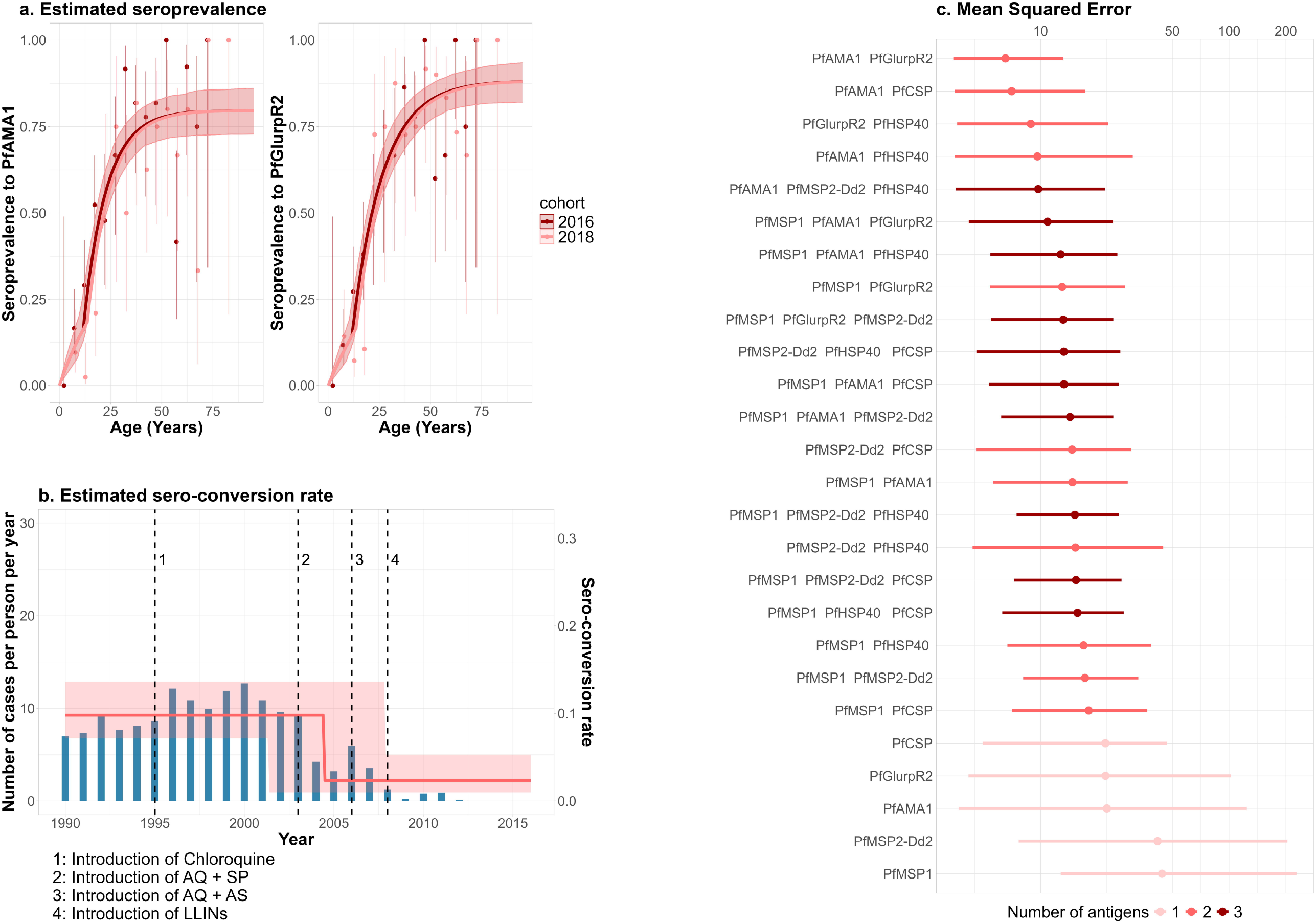
Antigens performance to determine seropositivity. Panel A show the distribution of PfCSP response compared to the bimodal distribution estimated by the GMM. Black vertical lines are the estimated means of the components representing the negatives and positives samples. Dashed coloured lines are the considered cutoffs. Panel B shows the analytical ROC curve from the GMM fitted on PfCSP as well as the different cutoffs considered and their associated sensibility and specificity. Panel C presents a ranking of antigens based on their performance to distinguish negative and positive samples as measured by AUC. Panel D shows the sensitivity associated with a cut-off that corresponding to a specificity of 99%. Estimates are shown as posterior medians with 95% credible intervals.

### Reconstruction of historical malaria transmission

A simple serocatalytic model including one change in transmission was first fitted to each antibody response observed in Ndiop where samples from children under five years old were available. Convergence issues for these simple serocatalytic models occurred for four antibody responses with low sensitivity (Erythrocyte Binding Antigen 4 (Etramp4.Ag2), *Pf*MSP-2.CH150, SBP1 and SEA1) (*Fig2*.*D*) and were ruled out of further analysis. Estimated sero-reversion rates were used as informative priors for models fitted to data from Dielmo. Models with up to three antigens were fitted to the Dielmo data. All models that converged were capable of recapturing age-stratified seroprevalence. All models estimated a sharp drop in transmission within the period of the four public health interventions with a decrease in transmission of greater than 50%. The best model, based on its ability to reconstruct the number of cases per person per year, includes both *Pf*AMA1 and *Pf*Glurp.R2, two long-lived blood stage antigens. *Pf*AMA1 is known to be very immunogenic and is recommended for serological surveillance in low transmission settings (20,21). Here, the estimated probability to seroconvert if exposed is 73% (56%, 88%) for *Pf*AMA1 and 60% (48%, 72%) for *Pf*Glurp.R2 (*Fig3*.*A*). This model estimated a drop in transmission in 2004, 11.6 (8.2, 14.6) years prior to 2016, with a sero-incidence rate of 0.10 (0.07, 0.14) year^-1^, before 2004 and a decrease of 76% (61%, 86%) afterwards (*Fig 3.B*). This coincides with the implementation of combination drugs (AQ+SP and AQ+AS) usage that were introduced systematically in Dielmo in 2003 and 2006, respectively. Models estimating a second drop in transmission were tested, but did not converge indicating that the data were not sufficiently informative to identify two changes in transmission.

**Fig 3.**
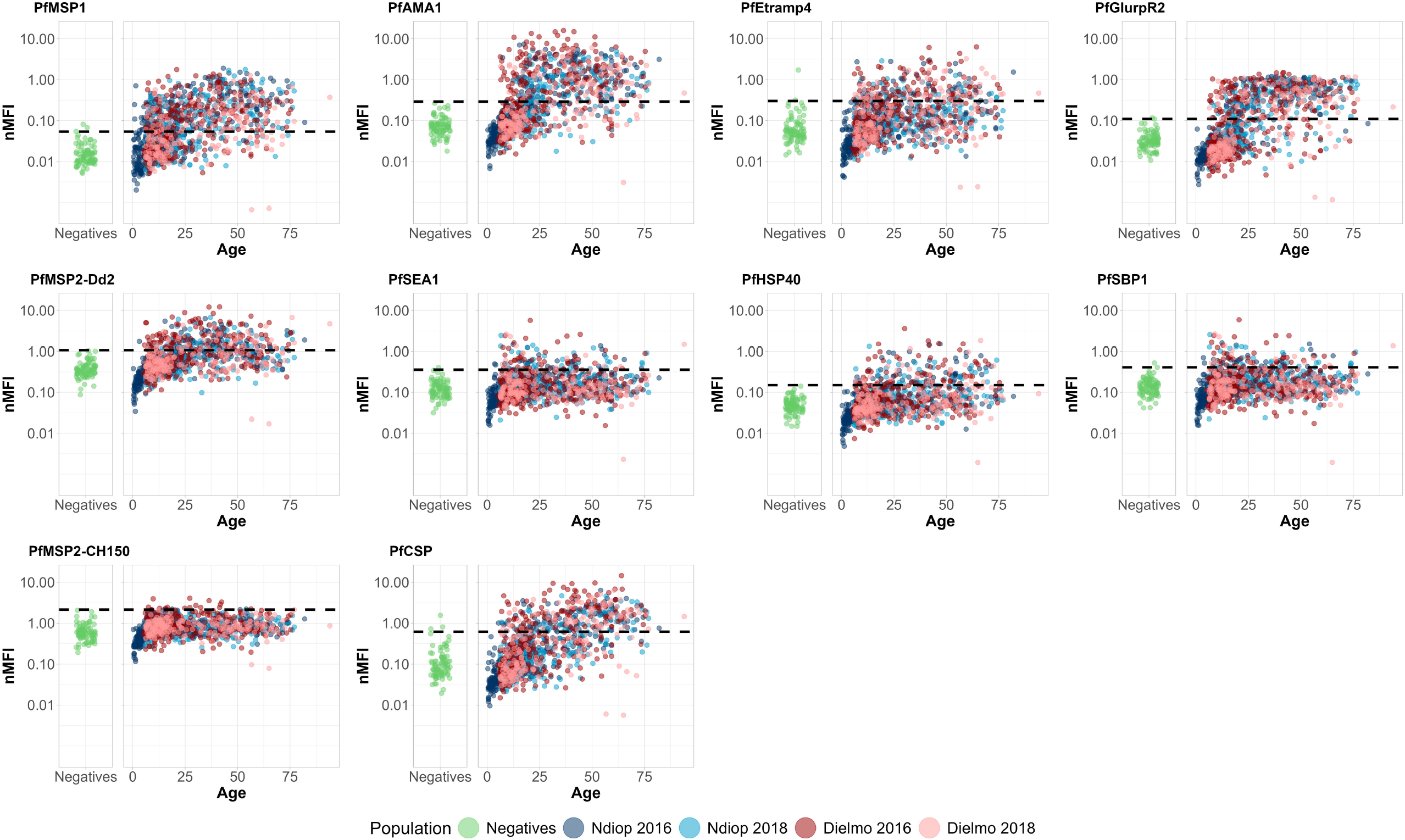
Model comparison to data and validation. Panel A compares Dielmo’s observed and model-predicted age-stratified seroprevalence, for a multiplex serocatalytic model including both PfAMA1 and PfGlurpR2 and considering one drop in transmission. In Panel B the sero-incidence estimated by the same model over 25 years prior to the 2016 cross section is compared to an external validation data on the observed number of cases per person per year. Panel C ranks every model that converged depending on their capability of recapturing the validation data. Mean Squared Error (MSE) between observation and model estimation and the associated 95%CI were computed after a conversion coefficient was applied to the posterior distribution of the sero-incidence. λ

### Multiplex sero-catalytic models provide more accurate estimation of transmission than singleplex models

The performance of models to reconstruct past transmission was validated against longitudinal data on cases of fever due to *P. falciparum* infection and the past implementation of public health interventions. The ranking of models based on mean squared error to this validation data differs statistically depending on the number of antigens included (Kruskal-Wallis p ≤ 0.01). The most accurate multiplex model (*Pf*AMA1 + *Pf*GLURP) outperformed all singleplex models. Overall, models including only one antigen provided less accurate estimates of past clinical incidence and show systematically higher MSE than models with multiple antigens. However, there were limited differences in MSE between models with two and three antigens. Not only are the models with one antigen less accurate at recapturing past transmission, but their uncertainty is wider as well (*Figure 3.C*). The same thing is found when looking at the posterior distributions of each parameter, singleplex serocatalytic models give less precise and more variable estimates, than multiplex models (*Supplementary Fig. S14-S23*).

## Discussion

In Dielmo, between 1990 and 2012 different public health interventions have been implemented reflecting Senegalese guidelines for *P. falciparum* case management. Following the development of parasite resistance to chloroquine, combination therapies were introduced, first amodiaquine plus sulfadoxine-pyrimethamine, and then amodiaquine plus artesunate (22). The impact of these new first line treatments, as well as the introduction of insecticide treated bed nets, can be observed in the reductions in parasite prevalence and case incidence in the longitudinal data collected over a 22 year period (22). It was also possible to reconstruct this reduction in transmission by fitting sero-catalytic models to measured antibody responses from cross-sectional surveys in 2016 and 2018. This study demonstrates the potential of multiplex serocatalytic models, as well as their limitations. While serocatalytic models fitted to ten antibody responses independently give similar estimates, synthetising those outputs into a unique measure of transmission is non-trivial. New models including two or three antibody responses have concordant estimates of the parameters. They also have less uncertainty in parameters estimation and are more reliable to reconstruct validation data.

Not all markers were capable of reconstructing historical *P. falciparum* transmission. Antibody responses to SBP1, SEA1 and *Pf*MSP-2.CH150 did not provide clear discrimination between exposed and unexposed individuals. Using them to quantify transmission with classification-based approaches may lead to unreliable estimates. Conversely, *Pf*Glurp.R2, PfCSP, *Pf*MSP1 and *Pf*AMA1 had wide dynamic ranges and were capable of discriminating exposed and unexposed populations. *Pf*AMA1 has been shown to be very immunogenic and relevant in a lot of different transmission settings (21,23). Combining *Pf*AMA1 with *Pf*Glurp.R2 correctly recaptured the 76% drop in transmission. However, uncertainty in the estimated time at which transmission drops is too wide to distinguish the different dates, two or three years apart, at which new treatments were implemented.

Another limitation of this study is that we were unable to use samples from children under 5 years from Dielmo. Since antibody acquisition reflects recent exposure, serological data from young participants are particularly informative in serocatalytic models. In this study, this was overcome by informing priors with the posterior distribution from a single antigen model fitted on Ndiop but it highlights the dependency of the model to information on young children which might be difficult to obtain.

This model takes advantage of the correlation between multiple antibody responses targeted at malaria, however if those are too correlated the information added to the model is not enough to estimate the additional parameters. Only 60% of models including more than one antibody response converged. This method might not be applicable in cases where information on only 2 or 3 antibody responses is available.

This modelling study takes place in a broader aim to leverage multiplex serological data now available, especially as a new study highlights the relevance of serocatalytic models estimates for decision making (24), and to model antibody responses and the correlation between them. While model taking into account cross reactivity have been developed (25) to our knowledge none have been implemented to look at multiple antibody responses to the same pathogen to determine the Force Of Infection.

In this study those new models for serological surveillance have been applied to inform Malaria transmission in an endemic region in Senegal, however this methodology could also be used on a wide variety of pathogens (arboviruses but also respiratory viruses, enteroviruses…) and, like serocatalytic models they can be applied to population in a broad variety of transmission settings.

## Materials and Methods

### A. Data

#### Serological assay

The antigens used in this study spanned the whole life cycle of *P. falciparum*, both blood and liver stage. All antigens were generated and expressed by *E. Coli* except for *Pf*AMA1 which was expressed in *Pichia Pastoris*. They were selected for their correlation to infections among children (11) or exposed volunteers (26) and their use was validated for malaria surveillance (11,12,26). Circumsporozoite protein (CSP) is present on sporozoites and is the antigen included in the WHO recommended RTS,S/AS01 and R21/Matrix-M malaria vaccines. Apical Membrane Antigen 1 (AMA1) is expressed on blood-stage merozoites, and to a lesser degree on sporozoites. Merozoite Surface Protein 1 (MSP1), Glutamate Rich Protein C-terminal repetitive segment (GLURP.R2), and the two major allelic types of Merozoite Surface Protein 2 (MSP-2.Dd2 and MSP-2.CH150), are targeted at merozoites’ surface. Etramp4.Ag2, HSP40.Ag1, SBP1 and SEA1 are targeted in infected red blood cell. A summary of these antigens’ characteristics and details of their development and production are reported in van den Hoogen *et al* (26).

The ten antigens of interest were coupled to Luminex microspheres to develop a multiplex bead-based assay (27)[Garcia et al. in preparation]. Briefly, using a KingFisher Duo Prime magnetic particle processor, a panel of 200 antigens including those ten were coupled to bead regions for identification. They were then incubated with the participant sera diluted at 1/400. After 3 washes, those complexes were incubated with a secondary antigen conjugated to R-phycoerythrin specific to IgG (Jackson Immunoresearch, UK) for quantification. After another round of washing and resuspension, samples were analysed by the Intelliflex system and results were given in Median Fluorescence Intensity (MFI). To take into account inter plate variability, MFI were standardised by being divided by the MFI value of a 1/400 dilution of a pool of sera exposed to malaria analysed on the same plate. These Normalized MFI (nMFI) were then used for analysis.

#### Samples

Dielmo is a village of about 400 inhabitants in western Senegal, north of the Gambian border, about four hours’ drive from the capital Dakar. The village is composed of traditional houses built with mud walls and villagers are mostly settled agricultural farmers. Small herds of cattle are also present in the vicinity of habitations. Malaria transmission is perennial (22,28). The Sudanian savanna climate of this region combined with the vegetation of the marshy banks of the river Nema support the presence of mosquitoes all year long, mostly *An gambiae* and *An funestus*. The majority of malaria cases are caused by *P. falciparum*, with a small proportion caused by *P. malariae* and *P. ovale*. At the time of initiation of the project in Dielmo, malaria transmission was holoendemic and prevalence was extremely high (86% of children in 1989 (28)), but it dropped in the following decades (<1% of children in 2012 (22)). Another village, Ndiop, was included in the study a few years later. Located a few kilometres south, Ndiop is very similar to Dielmo apart from the fact that transmission is concentrated during the rainy season (end of June to mid-October) (22).

Cross sectional surveys to investigate the acquisition and maintenance of natural immunity to malaria have been routinely conducted as part of a prospective longitudinal study in Dielmo and Ndiop villages in Senegal over the past 30 years(29–33). Blood samples were collected via venous bleed or finger prick from all consenting participants. Plasma and red blood cell were stored at-20°C after separation by centrifugation. This study focuses on the cross sections from 2016 and 2018.

In parallel, a prospective longitudinal study conducted between 1990 and 2012 in Dielmo recorded all cases of fever due to *P. falciparum* infection. Results from this systematic recording were given in number of cases per person per year. The treatment administered in case of infection depended on the national guidelines at the time and long lasting insecticides treated bed nets were introduced in the population during this period.

In 2020, a cohort was conducted in Oise (France) to study seroprevalence to SARS-CoV-2. Children and adults (n=1,132) attending hospital were recruited and blood samples drawn for routine medical care have been used in previous study for serological surveillance (27). In this study, a subset of 84 randomly selected samples were used as negative controls as participants are considered to not have been exposed to *P. falciparum*. Only adults were part of this sub sample with a median age of 53 [21, 90] years old, 44% of them were males.

#### Ethical consideration

The Senegalese samples were collected as part of the ongoing Dielmo and Ndiop project which was examined and approved by the Senegalese National Health Research Ethics Committee (CNERS Sénégal). Approval to measure antibodies to malaria in these samples was granted by CNERS Sénégal (N 00000007 MSAS/CNERS/Sec). Written informed consent was given by participants and parents or guardian of children enrolled.

The French cohort was approved by the Comité de Protection des Personnes Nord Ouest IV (NCT04644159) and samples were processed in line with the Commission Nationale de l’Informatique et des Libertes reglementation.

### B. Modelling

#### Sero-positivity classification

As serocatalytic models are fitted to seroprevalence data, continuous antibody levels measured in nMFI must be dichotomized as seropositive or seronegative to each antigen. A Gaussian Mixture Model was fitted to data on measured antibody responses, assuming that antibody levels in a population are a combination of two log normal distributions (one for the positive samples and one for the negatives). Here, Senegalese samples from both villages, whose serostatus is unknown, and French negative controls were included in the model allowing to account for all available information. Having negative controls was also helpful to inform the model in cases where the bimodal pattern was not clearly defined in the Senegalese data.

Overall, the data can be described as such:

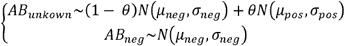

where *AB*_*unkown*_ is the measured antibody response of Senegalese samples, *AB*_*neg*_ the one of French negative controls, θ the proportion of positive samples in the Senegalese population and *µ*_*neg*_, *σ*_*neg*_, *µ*_*pos, σpos*_ the mean and standard deviation of the distribution of the antibody levels of the negative and positive samples respectively.

The estimated distributions were subjected to a Receiver Operating Characteristics (ROC) analysis to assess the trade-off between sensitivity and specificity. Sensitivity was estimated as the proportion of the CDF for the positive distribution higher than a selected cut-off, and specificity as the proportion of the CDF for the negative distribution below the selected cut-off. Three cut-offs were considered: one corresponding to the Youden Index, one corresponding to the estimated mean + 2*sd and one corresponding to a specificity of 99% but the later was preferred to the others that had too much variability in sensitivity and specificity between antigens. Calculating an analytical ROC curve based on the cumulative distribution functions of the estimated negative and positive distributions has been shown to have very similar results as an empirical ROC curve built on observed negative and positives samples (Yman et al. in preparation).

#### Serocatalytic model

Once the data is dichotomized, it is possible to reconstruct historic exposure by studying the age-stratified seroprevalence in this population. If we assume constant seroreversion rate *λ* and seroconversion rate *p* the proportion of a population that are seropositive at age *a* can be described by the following ODE system:

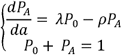

where *P*_*A*_ and *P*_0_ are the probabilities to be seropositive to antigen A and seronegative respectively at a given age. These equations can be solved analytically to give 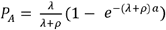 To study the impact of different public health interventions, two additional parameters were included: *tc* the time at which a sharp drop in transmission occurred and 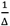 the magnitude of this drop. At a given age, the contribution to the likelihood is modeled as 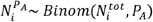

To simultaneously analyse data from antibody responses to multiple antigens, we extended existing sero-catalytic models. A model of the seroprevalence of two antigens can be described by the following system of ODEs:

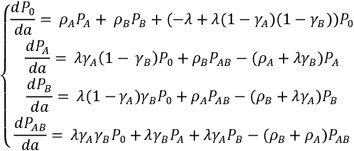

We assume a common serological incidence λ for all antibody responses. New parameters *γ*are included to estimate the probability of seroconversion if exposed. The seroconversion rate for antibody *A* is thus *λ γ*_*A*_

The equations for a model with three antigens as well as schema of the compartmental models are provided in the supplementary appendix. In the same way as in the model with a single antigen, the age-specific contribution to the likelihood is modelled using a multinomial distribution over serostatus. Once models were defined, a simulation/recapture study was conducted to assess the capacity of the model and inference framework to correctly estimate each parameter (*Supplementary III. b*.).

With each antigen added, the number of serostatus, and so the number of equations in the system increases exponentially while the number of parameters increases linearly. Even so this suggest that adding more antigens would not cause identifiability issues, the complexity of the models and computation time would become limiting. Meanwhile the information gained for each antigen added is decreasing due to high correlation between antibody responses. For those reasons, a maximum of three antibody responses were included simultaneously in the model.

In this study, data from two cross sections are available, one in 2016 and one in 2018. They were modelled simultaneously, with the same transmission pattern shifted by 2 years.

#### Implementation

Models were implemented in a Bayesian framework using Stan (version 2.32.2). Posterior distributions of the estimated parameters were obtained via Markov Chain Monte Carlo (MCMC) sampling, with four parallel chains run for 4,000 iterations each, including 1,000 warm-up iterations. Convergence was assessed for each model by graphical assessment of the mixing of the chains and posterior distributions. Visual predictive checks were used as well to ensure the adequacy of the model to the data.

ODE systems were solved using the ode_rk45 function in stan. Age was binned to one year for computational efficiency.

Most priors were as weakly informative as possible while being relevant: *tc ∼uniform(0,50), γ ∼Beta(3,1*.*3)* and *γ ∼exponential(1)*. For *ρ*, priors are specific to each antibody response. As stated previously, samples from children under five years old from Dielmo could not be analysed. Yet in serocatalytic models, *γ* and *ρ* are very correlated and are mostly informed by the younger age data which caused convergence issues. *ρ* is antigen dependent only and should be concordant in two similar populations. The choice was thus made to fit a simple serocatalytic model including one sharp drop in transmission to N’Diop data and use the posterior distribution as prior for.*ρ*

#### Protocol

Once the priors for *ρ* were described from a simple serocatalytic model, they were applied to all combination of 1, 2 or 3 antibody responses observed in Dielmo out of the 6 that showed no convergence issues in N’Diop. Models that converged were considered for external validation. Finally, a Kruskall Wallis ranking test was conducted to assess the importance of taking into account more than one antibody response at a time (*Supplementary III*.*c*).

#### External validation

Data from figure 3.C of (22), was reported using a pixel measurer and used as external validation for parameters estimated from the different models. As *λ* is not equivalent and can not be converted to number of cases per participant per year, a conversion coefficient was computed that minimizes the mean standard error (MSE) between median parameters and the data. This coefficient was then used on the whole range of the posterior distribution to recapture uncertainty. MSE computed from the median and IC95% were then compared to select the best model to recapture the past implementation of public health interventions. In this framework, only *tc* and Δ are validated as *λ* has been converted and *ρ* can not be recaptured by the validation data.

## Supporting information

Supplementary methods and results

## Data Availability

All data produced are available online at https://doi.org/10.5281/zenodo.17368372

## Acknowledgments

This work was supported by the European Research Council (MultiSeroSurv, 852373), and the French government’s “Integrative Biology of Emerging Infectious Diseases” (Investissement d’Avenir grant ANR-10-LABX-62-IBEID) and INCEPTION programs (Investissement d’Avenir grant ANR-16-CONV-0005). The authors thank the study participants and community health workers in Dielmo and Ndiop villages.

## Supporting Information

### S1 Text. Supplementary information on methods and results

Antigen descriptions, schematic representations of multiplexed models, assessment of the inference framework, protocol flowchart and systematic display of modelling results.

## Notes

**Data availability statement:** All relevant data are within the manuscript and its Supporting Information files.

### Competing Interest Statement

The authors have declared no competing interest.

### Clinical Trial

NCT04644159

### Funding Statement

This study was funded by the European Research Council (MultiSeroSurv, 852373) and the French government s Integrative Biology of Emerging Infectious Diseases (Investissement d Avenir grant ANR-10-LABX-62-IBEID) and INCEPTION programs (Investissement d Avenir grant ANR-16-CONV-0005)

### Author Declarations

Ethics committee of Senegalese National Health Research (CNERS Senegal) gave ethical approval for this work (N 00000007 MSAS/CNERS/Sec). Ethics committee of Comite de Protection des Personnes Nord Ouest IV gave ethical approval for this work (NCT04644159).

