## Supplementary methods and results for "Reconstruction of historical malaria transmission in Senegal using multiplex sero-catalytic models"

Supporting Information

I. Antigens

| **Name** | **Description** | **Location** | **Expression tag** |
| --- | --- | --- | --- |
| ***PfCSP*** | Circumsporozoite protein | Sporozoites | GST |
| ***PfGlurpR2*** | Glutamate Rich Protein C-terminal repetitive segment | Merozoites surface | GST |
| ***PfAMA1*** | Apical Membrane Antigen 1 | Blood stage merozoites / sporozoites | His |
| ***PfMSP1*** | Merozoite Surface Protein 1 | Merozoites surface | GST |
| ***PfEtramp4*** | Early Transcribed Membrane Protein 4 | Infected red blood cell | GST |
| ***PfHSP40*** | Heat Shock Protein 40 | Infected red blood cell | GST |
| ***PfMSP2-Dd2*** | Merozoite Surface Protein 2 (Dd2 allele) | Merozoites surface | GST |
| ***PfMSP2-CH150*** | Merozoite Surface Protein 2 (CH150 allele) | Merozoites surface | GST |
| ***PfSEA1*** | Schizont Egress Antigen 1 | Infected red blood cell | GST |
| ***PfSBP1*** | Skeleton Binding Protein 1 | Infected red blood cell | GST |

***Table S1: Antigens description.*** *Characteristics of the 10 antigens analysed in this study.*

II. Classification

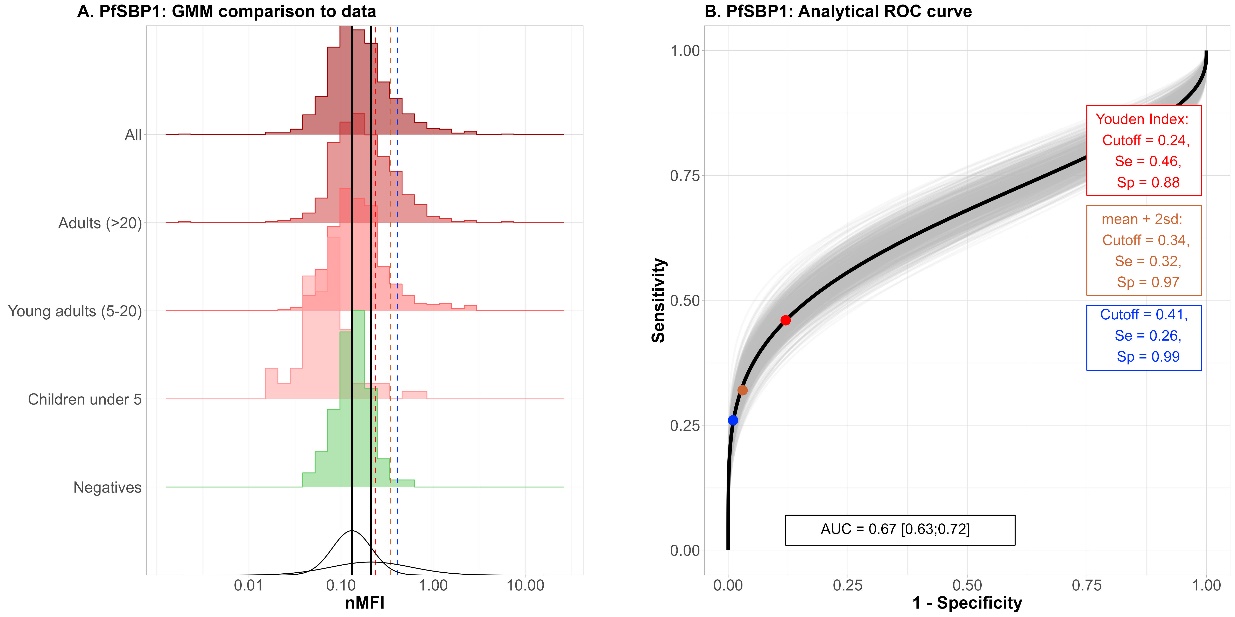

***Figure S1: GMM fitted to PfSBP1.*** *Panel A shows the distribution of PfSBP1 response compared to the bimodal distribution estimated by the GMM. Black vertical lines are the estimated means of the components representing the negatives and positives samples. Dashed coloured lines are the considered cutoffs. Panel B shows the analytical ROC curve from the GMM fitted on PfSBP1 as well as the different cutoffs considered and their associate sensibility, specificity.*

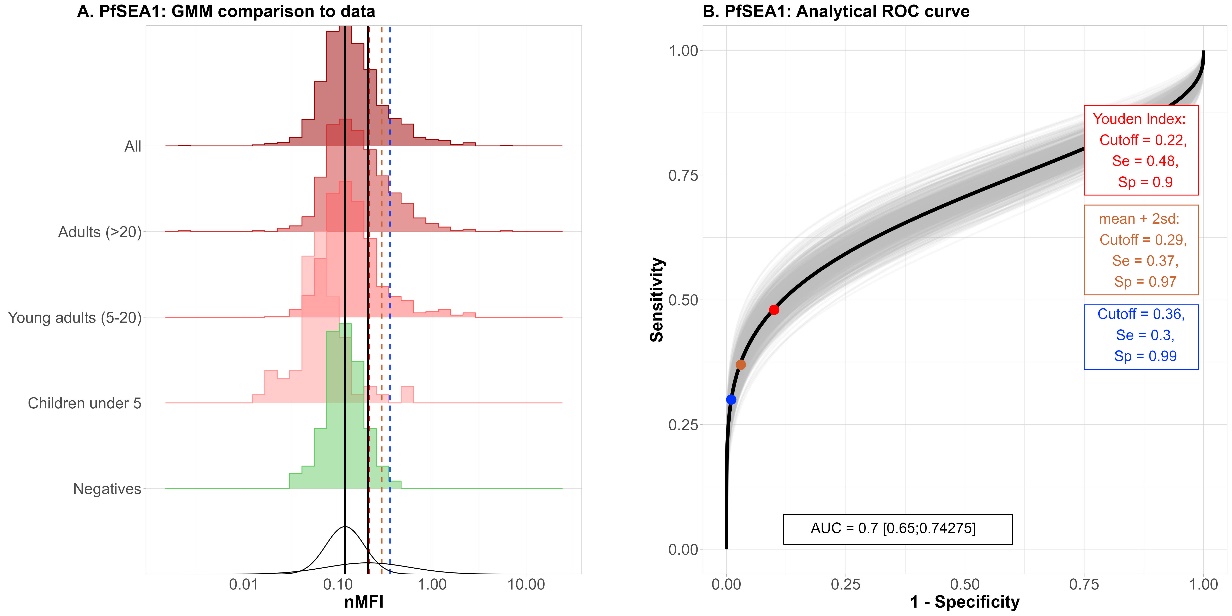

***Figure S2: GMM fitted to PfSEA1.*** *Panel A shows the distribution of PfSEA1 response compared to the bimodal distribution estimated by the GMM. Black vertical lines are the estimated means of the components representing the negatives and positives samples. Dashed coloured lines are the considered cutoffs. Panel B shows the analytical ROC curve from the GMM fitted on PfSEA1 as well as the different cutoffs considered and their associate sensibility, specificity.*

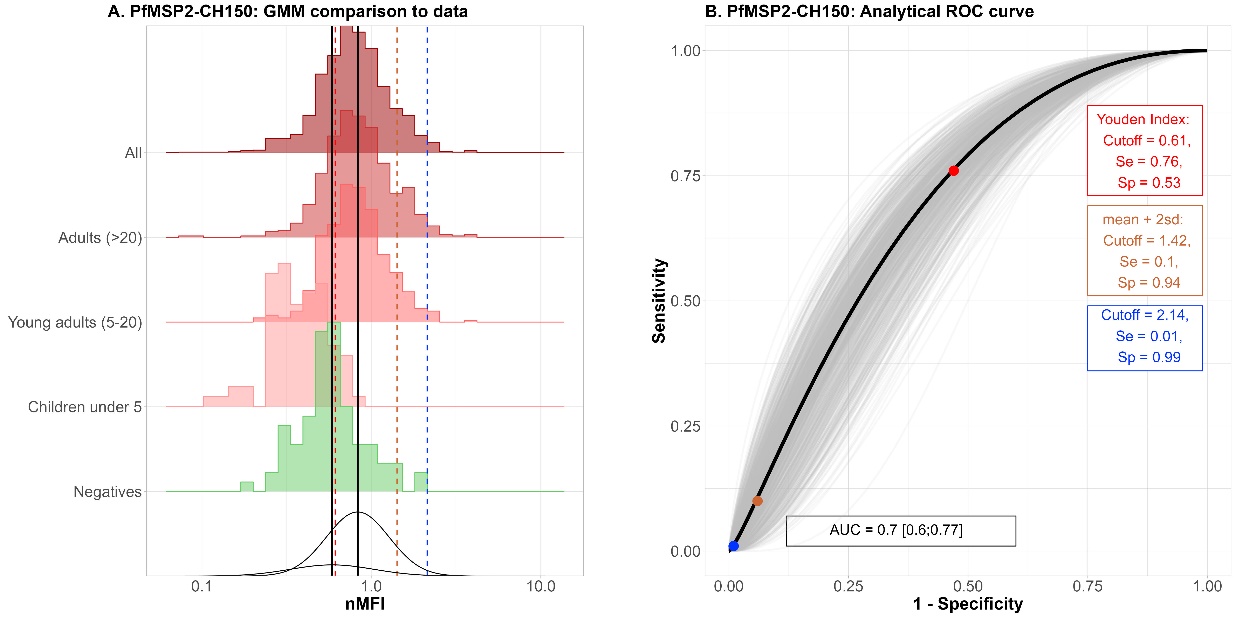

***Figure S3: GMM fitted to PfMSP2-CH150.*** *Panel A shows the distribution of PfMSP2-CH150 response compared to the bimodal distribution estimated by the GMM. Black vertical lines are the estimated means of the components representing the negatives and positives samples. Dashed coloured lines are the considered cutoffs. Panel B shows the analytical ROC curve from the GMM fitted on PfMSP2-CH150 as well as the different cutoffs considered and their associate sensibility, specificity.*

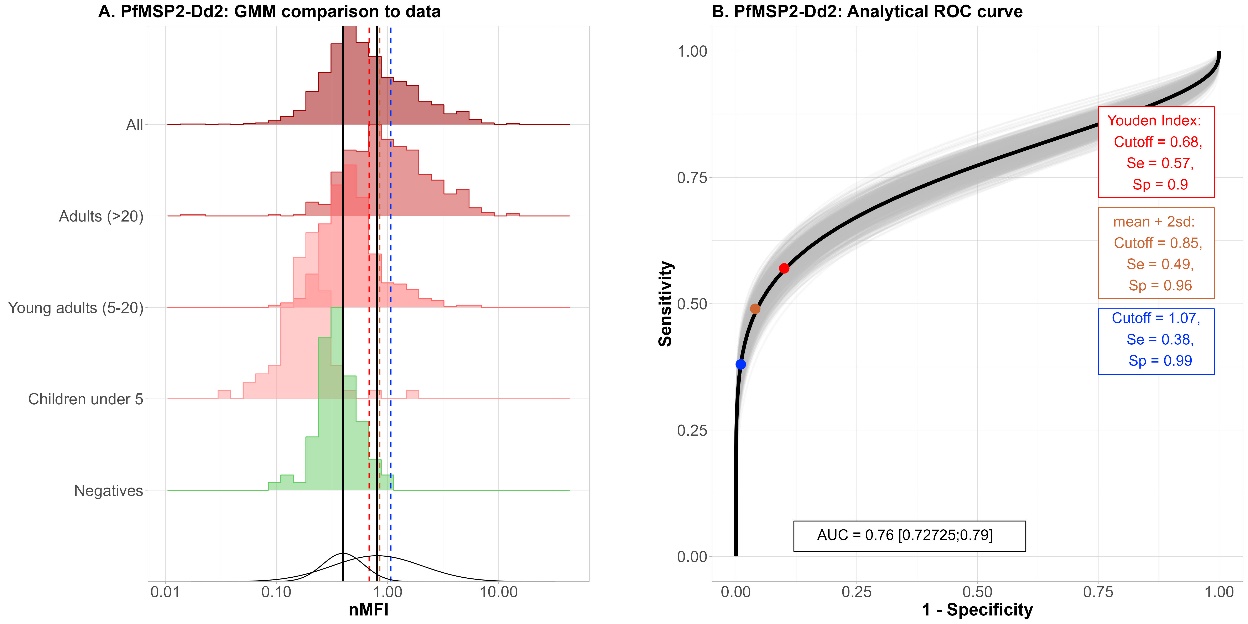

***Figure S4: GMM fitted to PfMSP2-Dd2.*** *Panel A shows the distribution of PfMSP2-Dd2 response compared to the bimodal distribution estimated by the GMM. Black vertical lines are the estimated means of the components representing the negatives and positives samples. Dashed coloured lines are the considered cutoffs. Panel B shows the analytical ROC curve from the GMM fitted on PfMSP2-Dd2 as well as the different cutoffs considered and their associate sensibility, specificity.*

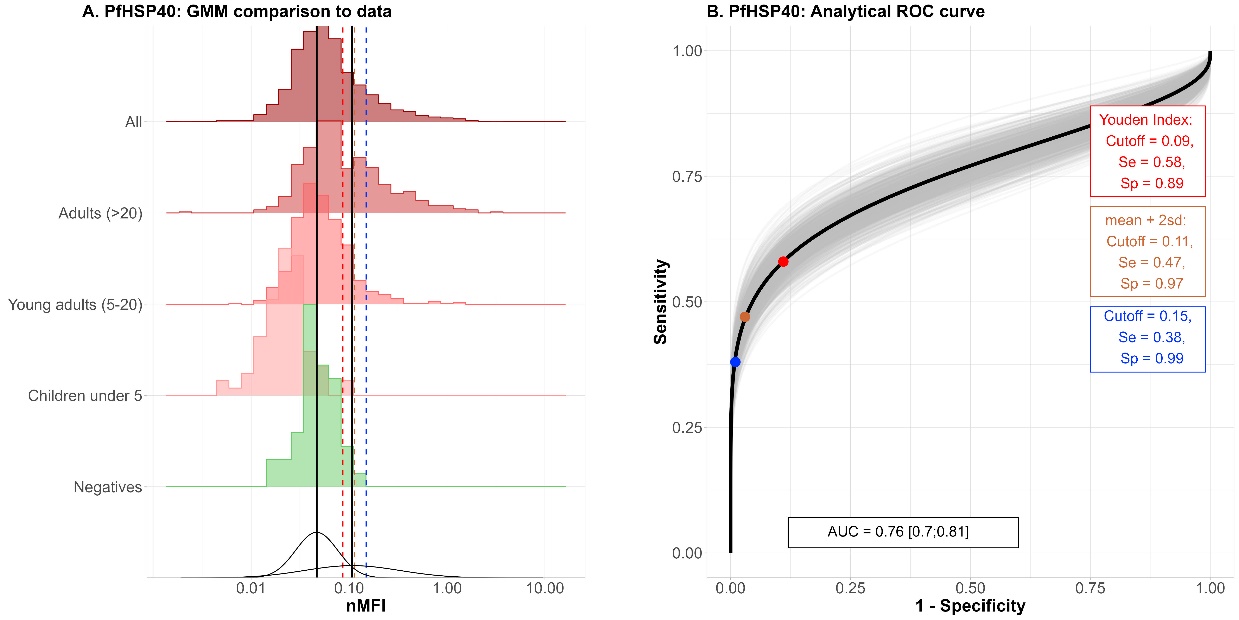

***Figure S5: GMM fitted to PfHSP40.*** *Panel A shows the distribution of PfHSP40 response compared to the bimodal distribution estimated by the GMM. Black vertical lines are the estimated means of the components representing the negatives and positives samples. Dashed coloured lines are the considered cutoffs. Panel B shows the analytical ROC curve from the GMM fitted on PfHSP40 as well as the different cutoffs considered and their associate sensibility, specificity.*

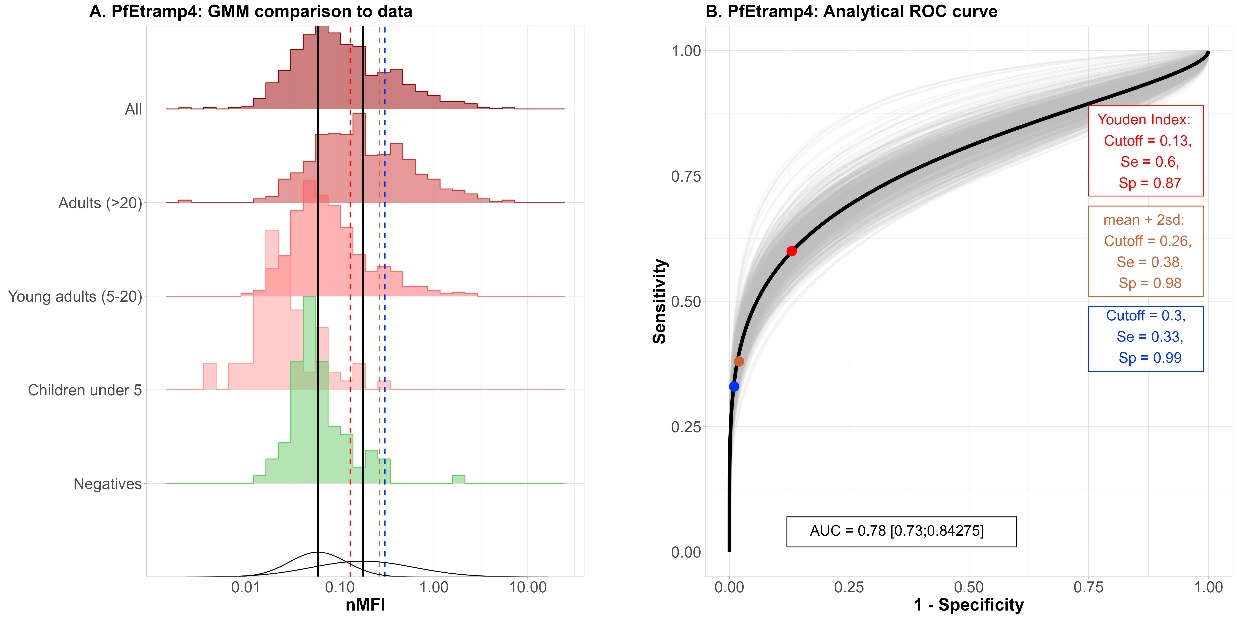

***Figure S6: GMM fitted to PfEtramp4.*** *Panel A shows the distribution of PfEtramp4 response compared to the bimodal distribution estimated by the GMM. Black vertical lines are the estimated means of the components representing the negatives and positives samples. Dashed coloured lines are the considered cutoffs. Panel B shows the analytical ROC curve from the GMM fitted on PfEtramp4 as well as the different cutoffs considered and their associate sensibility, specificity.*

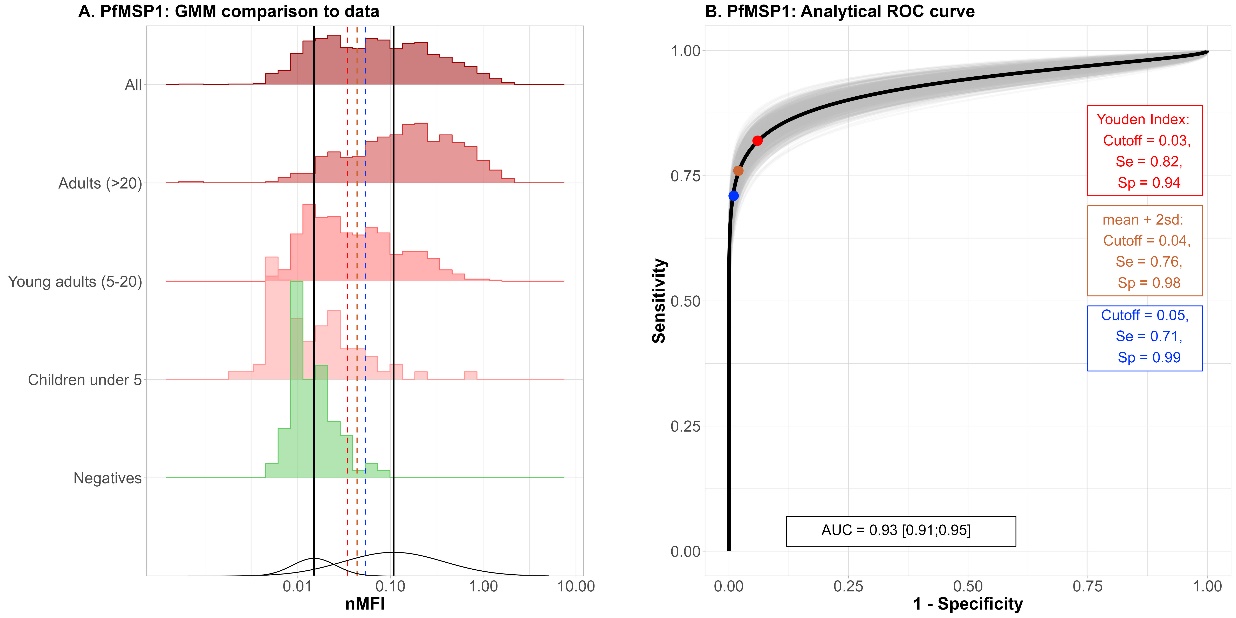

***Figure S7: GMM fitted to PfMSP1.*** *Panel A shows the distribution of PfMSP1 response compared to the bimodal distribution estimated by the GMM. Black vertical lines are the estimated means of the components representing the negatives and positives samples. Dashed coloured lines are the considered cutoffs. Panel B shows the analytical ROC curve from the GMM fitted on PfMSP1 as well as the different cutoffs considered and their associate sensibility, specificity.*

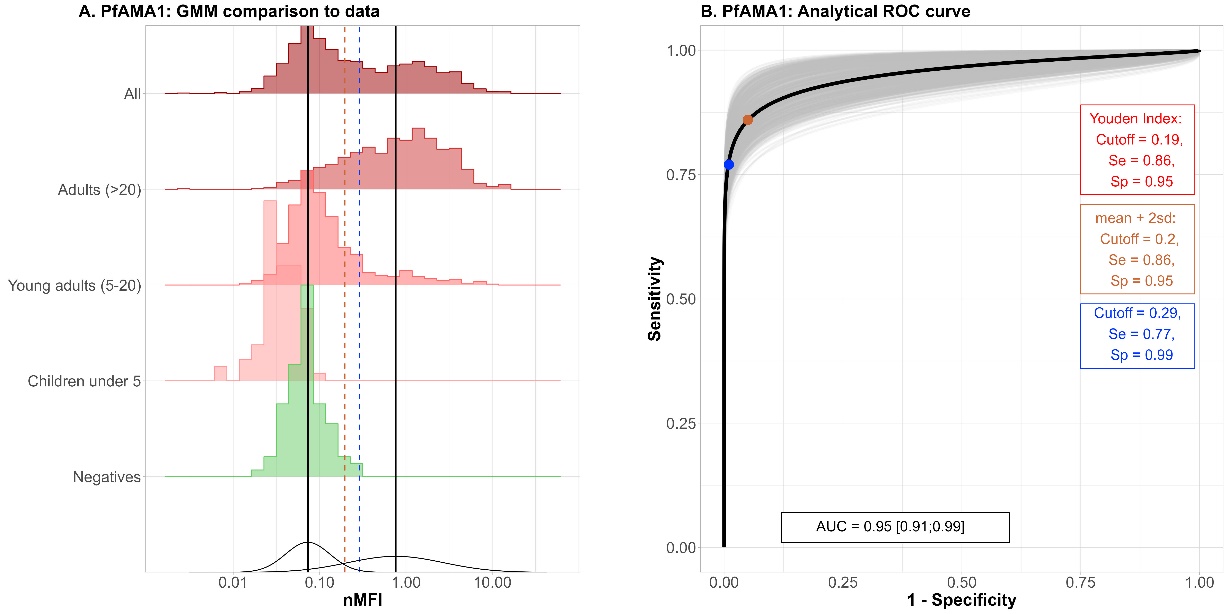

***Figure S8: GMM fitted to PfAMA1.*** *Panel A shows the distribution of PfAMA1 response compared to the bimodal distribution estimated by the GMM. Black vertical lines are the estimated means of the components representing the negatives and positives samples. Dashed coloured lines are the considered cutoffs. Panel B shows the analytical ROC curve from the GMM fitted on PfAMA1 as well as the different cutoffs considered and their associate sensibility, specificity.*

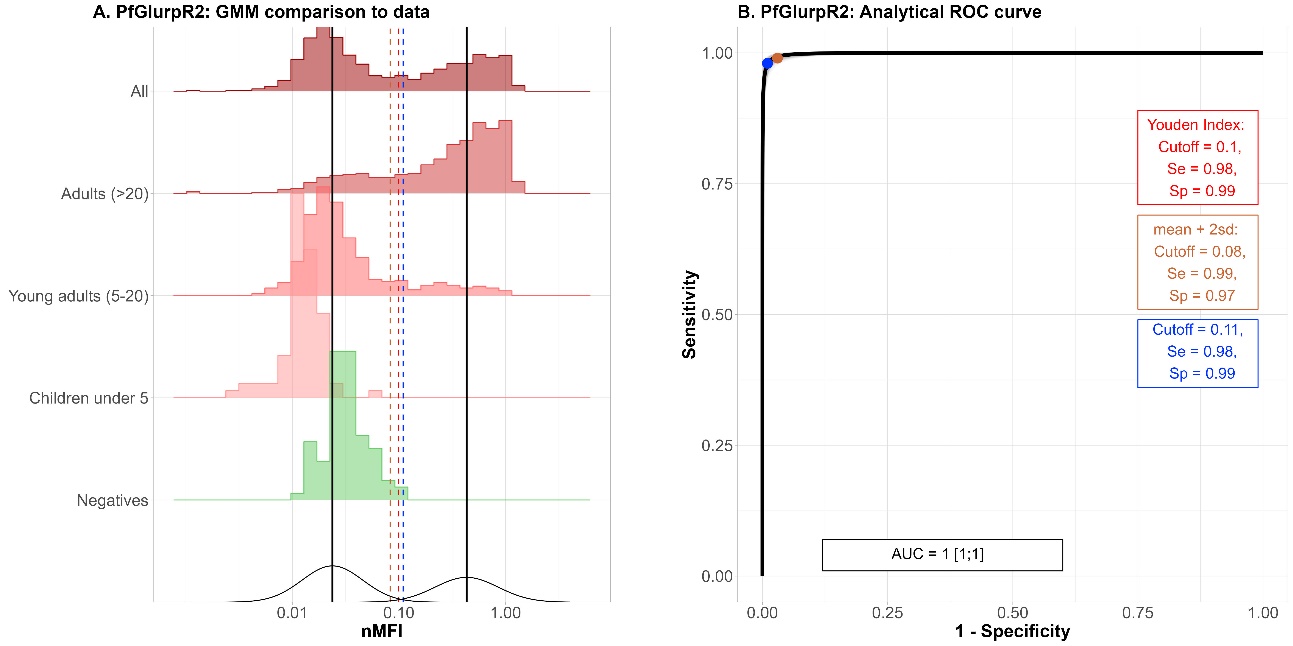

***Figure S9: GMM fitted to PfGlurpR2.*** *Panel A shows the distribution of PfGlurpR2 response compared to the bimodal distribution estimated by the GMM. Black vertical lines are the estimated means of the components representing the negatives and positives samples. Dashed coloured lines are the considered cutoffs. Panel B shows the analytical ROC curve from the GMM fitted on PfGlurpR2 as well as the different cutoffs considered and their associate sensibility, specificity.*

III. Modelling
     a) Model definition

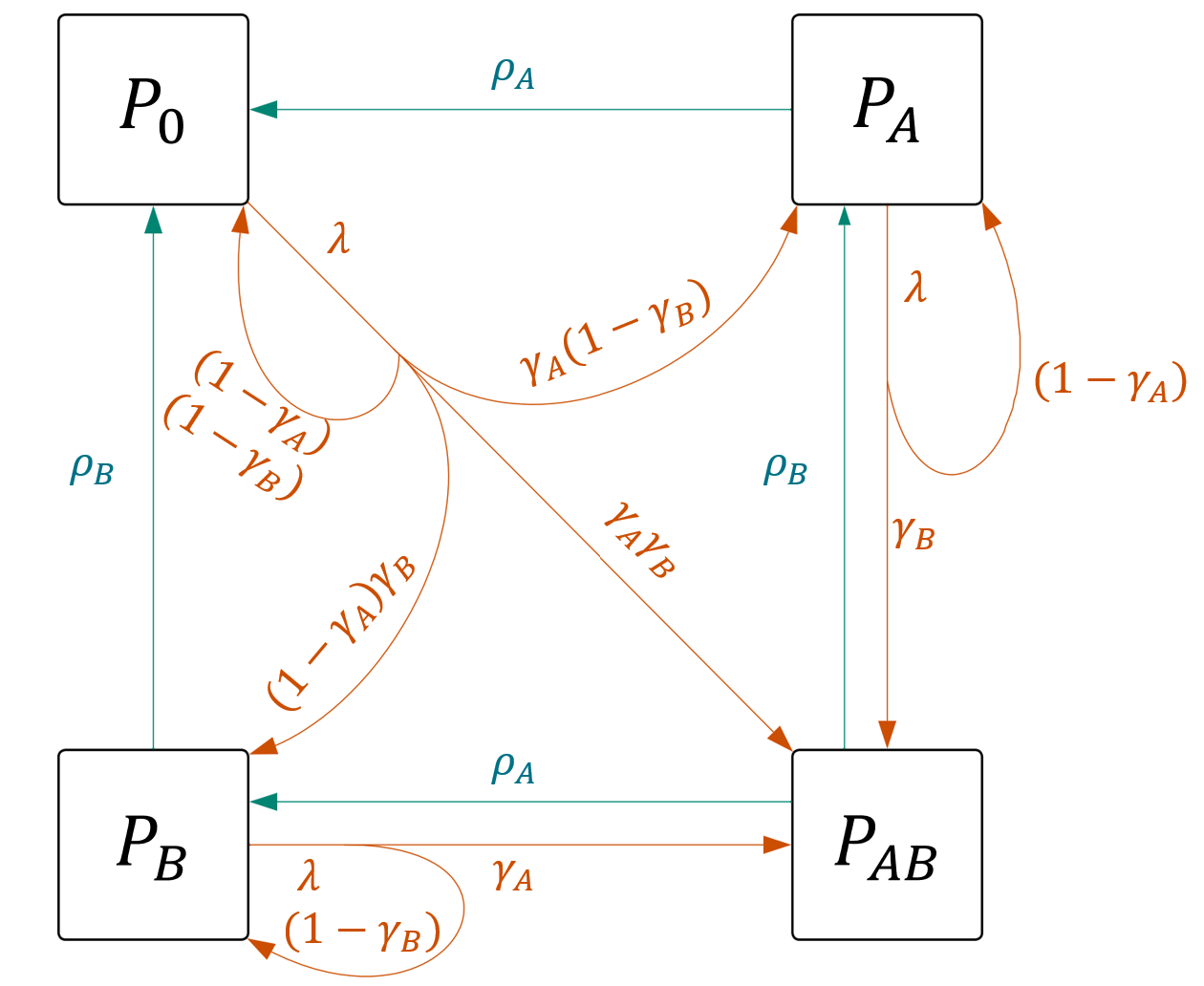

***Figure S10: Schematic representation of a serocatalytic model including two antibody responses.*** *Let A and B be two antibodies:* $P_{0}$ *is seronegative,* $P_{A}$ *is positive to A only,* $P_{B}$ *is positive to B only and* $P_{AB}$ *is positive to both. This model considers four possible serostatus, the serological incidence, the probability to seroconvert once exposed (red arrows) for each antibody and waning of each antibody (blue arrows).*

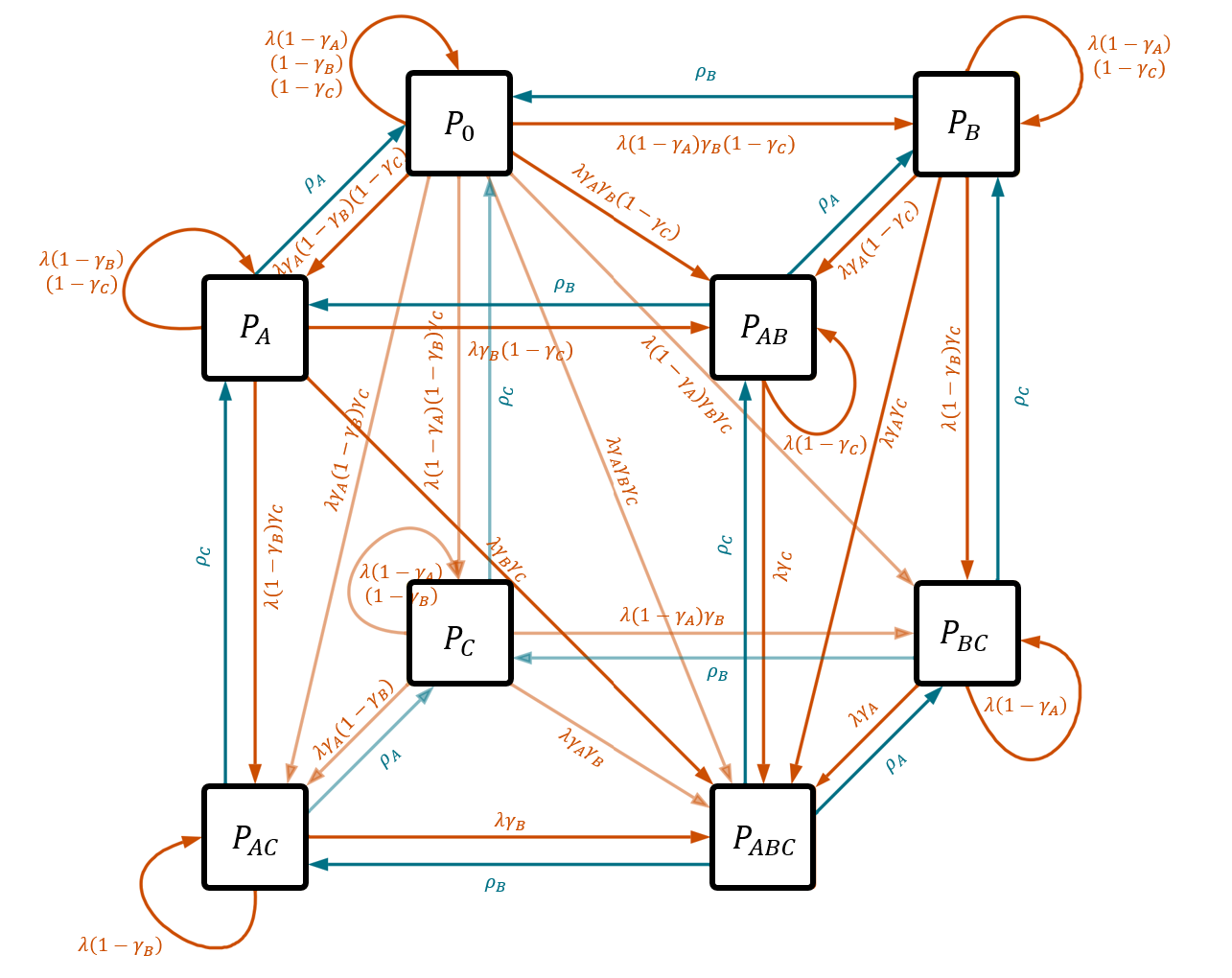

***Figure S11: Schematic representation of a serocatalytic model including three antibody responses.*** *Let A, B and C be three antibodies:* $P_{0}$ *is seronegative,* $P_{A}$*,* $P_{B}$ *and* $P_{C}$ *is positive only to A, B or C respectively,* $P_{AB}$*,* $P_{AC}$*,* $P_{BC}$ *are positive to each combination of two out of the three antibodies and* $P_{ABC}$ *is seropositive to A, B and C. This model considers eight possible serostatuses, the serological incidence, the probability to seroconvert once exposed (red arrows) for each antibody and waning of each antibody (blue arrows).*

$$\left\{ \begin{aligned} \frac{dP_{0}}{da}= \rho_{A}P_{A}+ \rho_{B}P_{B}+ \rho_{C}P_{C}+(-\lambda+\lambda(1-\gamma_{A})(1-\gamma_{B})(1-\gamma_{C}))P_{0} \\ \frac{dP_{A}}{da}= \lambda\gamma_{A}\left( 1- \gamma_{B} \right)\left( 1- \gamma_{C} \right)P_{0}+\rho_{B}P_{AB}+\rho_{C}P_{AC}+(-\lambda+\lambda\left( 1-\gamma_{B} \right)\left( 1-\gamma_{C} \right)-\rho_{A})P_{A} \\ \frac{dP_{B}}{da}= \lambda\left( 1-\gamma_{A} \right)\gamma_{B}\left( 1- \gamma_{C} \right)P_{0}+\rho_{A}P_{AB}+\rho_{C}P_{BC}+\left( -\lambda+\lambda\left( 1-\gamma_{A} \right)\left( 1-\gamma_{C} \right)-\rho_{B} \right)P_{B} \\ \frac{dP_{C}}{da}= \lambda(1-\gamma_{A})(1-\gamma_{B})\gamma_{C}P_{0}+\rho_{A}P_{AC}+\rho_{B}P_{BC}+(-\lambda+\lambda\left( 1-\gamma_{A} \right)\left( 1-\gamma_{B} \right)-\rho_{C})P_{C} \\ \frac{dP_{AB}}{da}= \lambda\gamma_{A}{(1-\gamma}_{C})P_{B}+\lambda\gamma_{B}{(1-\gamma}_{C})P_{A}+\rho_{C}P_{ABC}+(-\lambda+\lambda\left( 1-\gamma_{C} \right)-\rho_{B}-\rho_{A})P_{AB} \\ \frac{dP_{BC}}{da}= \lambda{(1-\gamma}_{A})\gamma_{C}P_{B}+\lambda{(1-\gamma}_{A})\gamma_{B}P_{C}+\rho_{A}P_{ABC}+(-\lambda+\lambda\left( 1-\gamma_{A} \right)-\rho_{B}-\rho_{C})P_{BC} \\ \frac{dP_{AC}}{da}= \lambda\gamma_{A}{(1-\gamma}_{B})P_{C}+\lambda{(1-\gamma}_{B})\gamma_{C}P_{A}+\rho_{B}P_{ABC}+(-\lambda+\lambda\left( 1-\gamma_{B} \right)-\rho_{A}-\rho_{C})P_{AC} \\ \frac{dP_{ABC}}{da}= \lambda\gamma_{A}P_{BC}+\lambda\gamma_{B}P_{AC}+\lambda\gamma_{C}P_{AB}+\lambda\gamma_{A}\gamma_{B}P_{C}+\lambda\gamma_{A}\gamma_{C}P_{B}+\lambda\gamma_{B}\gamma_{C}P_{A}+\lambda\gamma_{A}\gamma_{B}\gamma_{C}P_{0}-(\rho_{A}+\rho_{B}+\rho_{C})P_{ABC} \end{aligned} \right.$$

***Equation S1: ODE system of a model considering three antibody responses.*** $\lambda$ *is a common serological incidence,* $\gamma_{A}$*,* $\gamma_{B}$*,* $\gamma_{C}$ *are the probabilities to seroconvert if exposed for each antibody and* $\rho_{A}$*,* $\rho_{B}$*,* $\rho_{C}$ *the antibody dependent seroreversion rates.*

b) Simulation / Recapture study

Once models were defined, a simulation/recapture study was conducted to assess the capacity of the model and inference framework to correctly estimate each parameter. Populations were simulated assuming a Senegalese age distribution under different scenarios of malaria exposure. The serostatus attributed to each participant was sampled in a multinomial distribution with the probabilities computed by the model to be tested. For each set of parameters (*Table S2*), 20 populations were simulated and the model was fitted on each of them in a Bayesian framework using one Markov Chain of 800 iterations in R stan. The merged posterior distributions of the estimated parameters were then compared to their actual value used for the simulations. The validation of the model was made by visual assessment of those comparisons, plus the Visual Predictive Check (VPC) comparison to the simulated data.

| **Parameter** | **Values tested** |
| --- | --- |
| Serological incidence $\lambda$ (${years}^{-1}$) | 0.01, 0.1, 1 |
| Seroreversion rates $\rho$ (${years}^{-1}$) | 0.01, 0.1 |
| Probability to seroconvert if exposed $\gamma$ | 0.3, 0.8 |
| Time of drop in transmission $tc$ ($years$) | 5, 15 |
| Magnitude of drop in transmission $\frac{1}{\Delta}$ | 0.1, 0.5 |

***Table S2: Parameter values tested in the simulation recapture study.*** *The serocatalytic model including two antibody response and one sharp drop in transmission was fitted on 20 populations simulated for each combination of those parameter values.*

More than 80% (51 out of 62) of the models were capable of recapturing the parameters used for simulation (*Figure S12*).

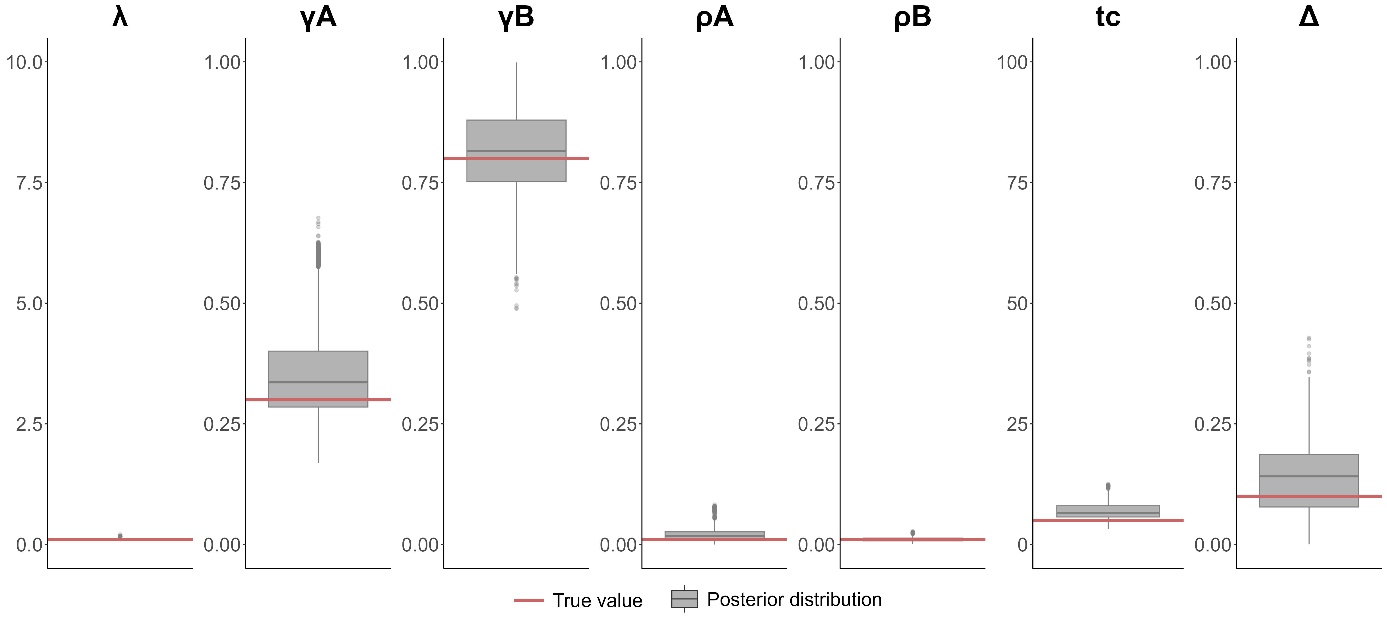

***Figure S12: Posterior distributions.*** *The model was fitted to 20 populations simulated with the sero-incidence* $\lambda=0.1$*, the probabilities to seroconvert to antigen A and B, respectively* $\gamma_{A}=0.3$ *and* $\gamma_{B}=0.8$*, the seroreversion rates* $\rho_{A}=0.01$ *and* $\rho_{B}=0.01$*, the time of the sharp drop in transmission* $tc=5$ *and a new sero-incidence* $\lambda\Delta=0.1*0.1$*. The grey boxplots are the merged posterior distributions of those 20 fits compared to the actual parameter values.*

c) Framework

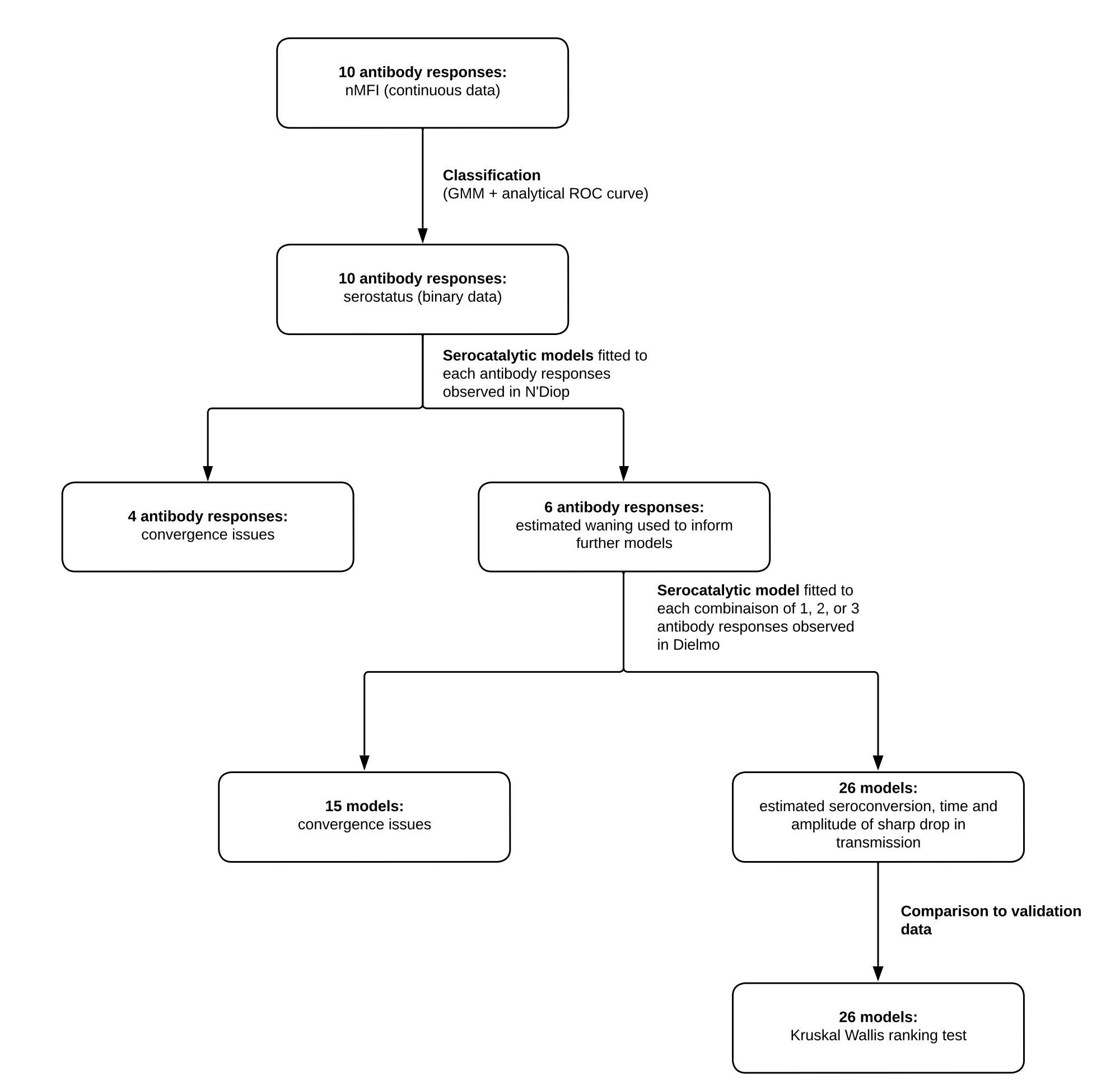

***Figure S13: Protocol flowchart.***

d) Posterior distributions

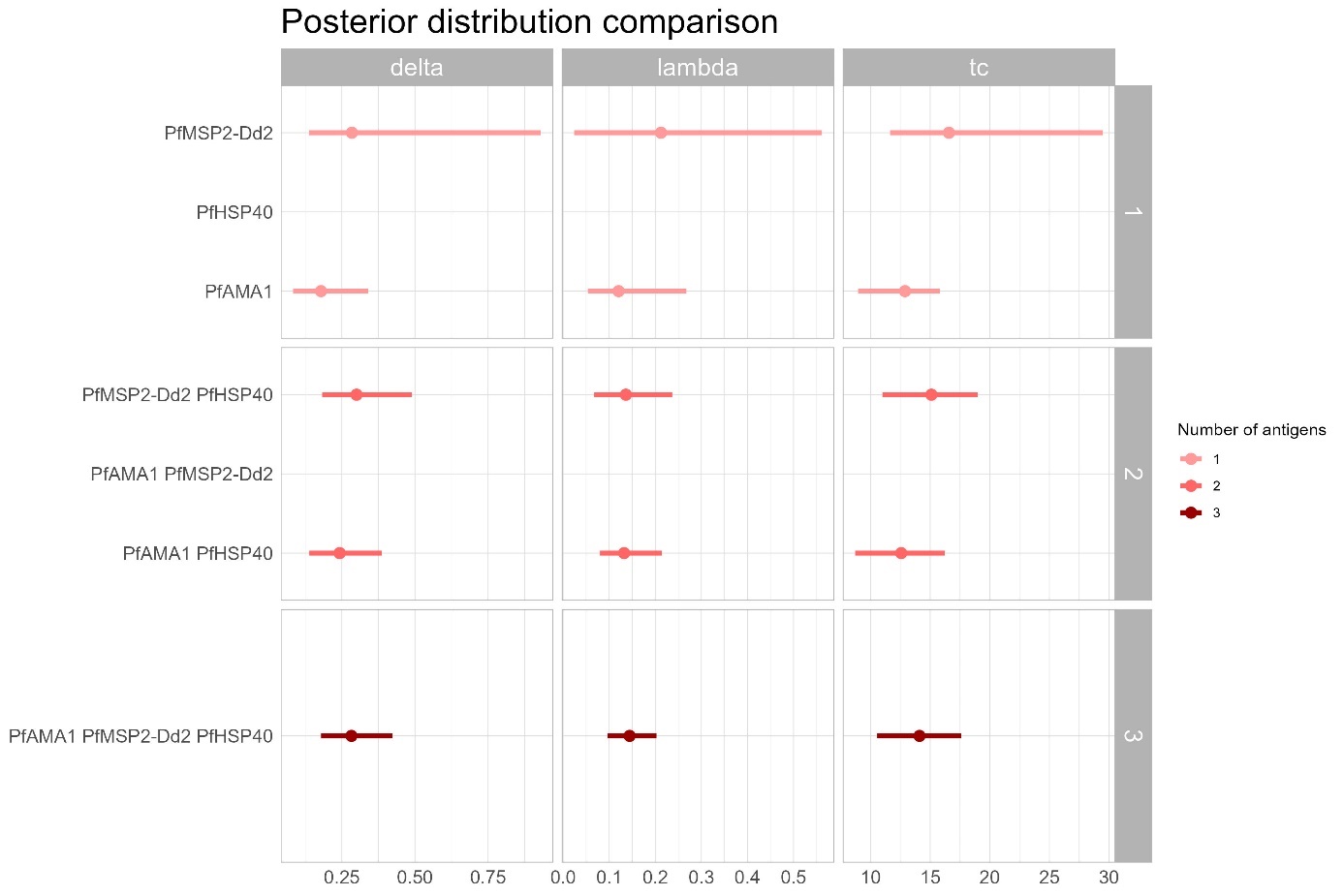

***Figure S14: Posterior distribution comparison.*** *Posterior distribution of the three parameters that have been compared to validation data: sero-incidence, time and magnitude of sharp drop in transmission. Model presented in this plot are all possible combinations of PfAMA1, PfMSP2-Dd2 and PfHSP40. Only models that converged are presented.*

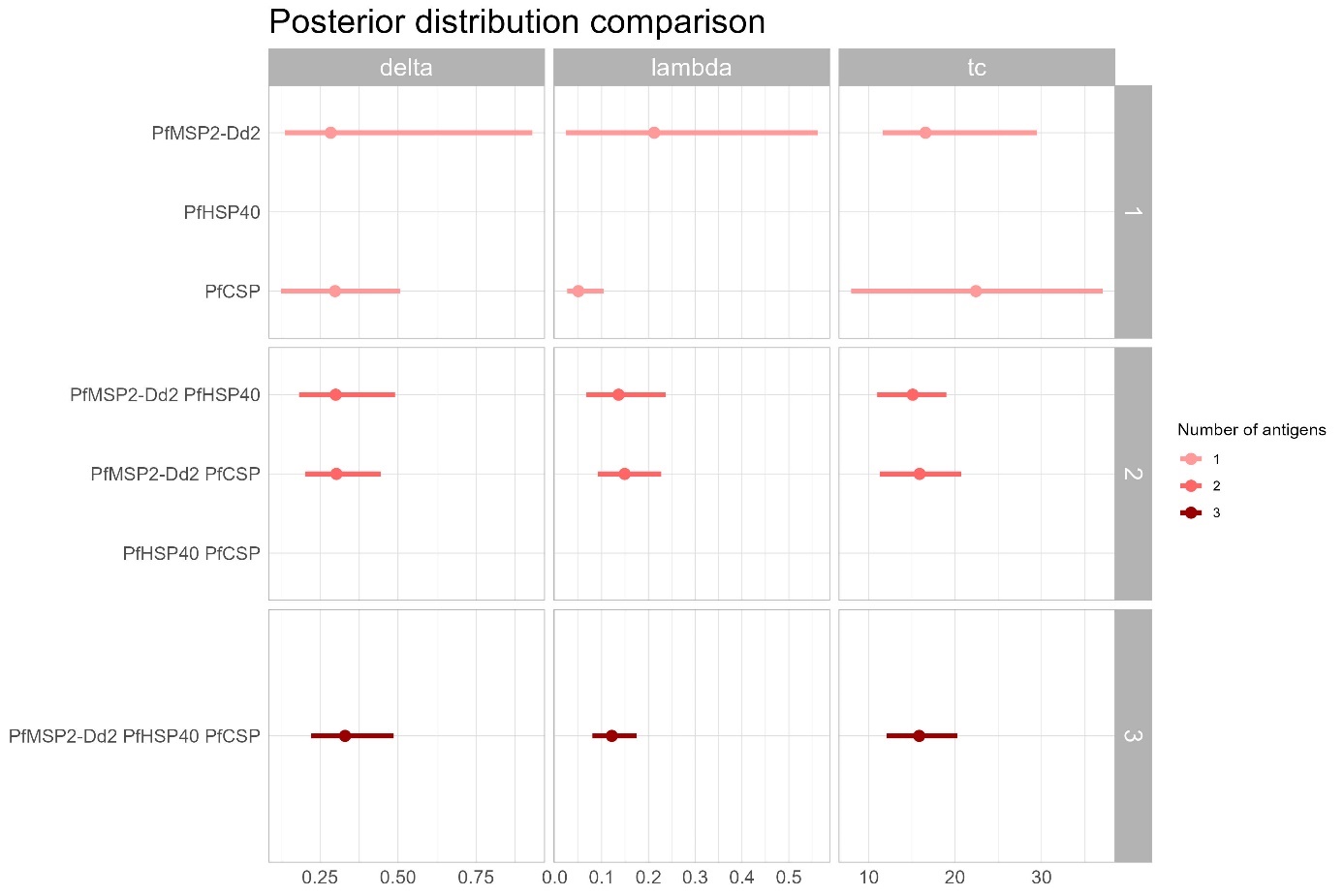

***Figure S15: Posterior distribution comparison.*** *Posterior distribution of the three parameters that have been compared to validation data: sero-incidence, time and magnitude of sharp drop in transmission. Model presented in this plot are all possible combinations of PfMSP2-Dd2, PfHSP40 and PfCSP. Only models that converged are presented.*

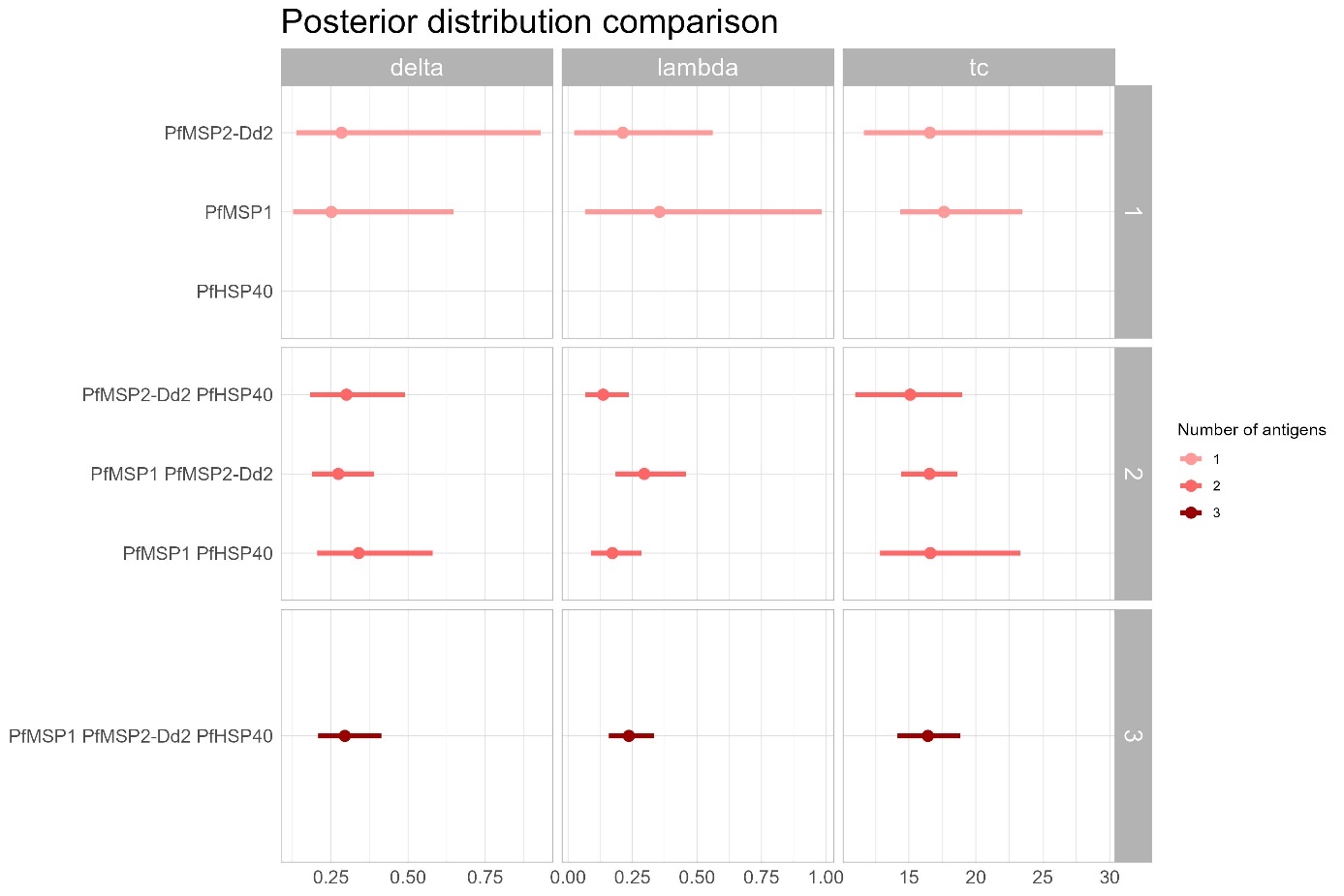

***Figure S16: Posterior distribution comparison.*** *Posterior distribution of the three parameters that have been compared to validation data: sero-incidence, time and magnitude of sharp drop in transmission. Model presented in this plot are all possible combinations of PfMSP1, PfMSP2-Dd2 and PfHSP40. Only models that converged are presented.*

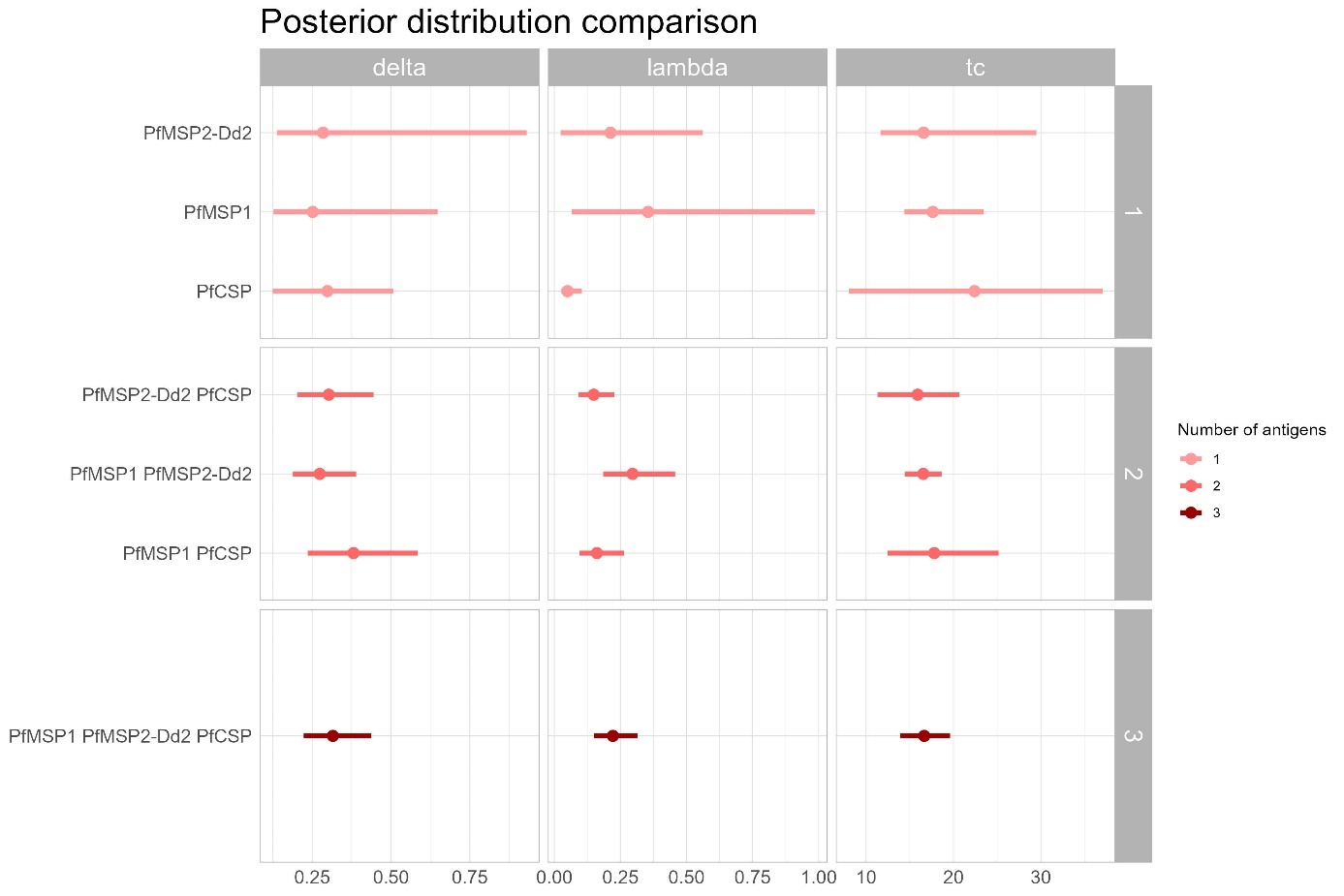

***Figure S17: Posterior distribution comparison.*** *Posterior distribution of the three parameters that have been compared to validation data: sero-incidence, time and magnitude of sharp drop in transmission. Model presented in this plot are all possible combinations of PfMSP1, PfMSP2-Dd2 and PfCSP. Only models that converged are presented.*

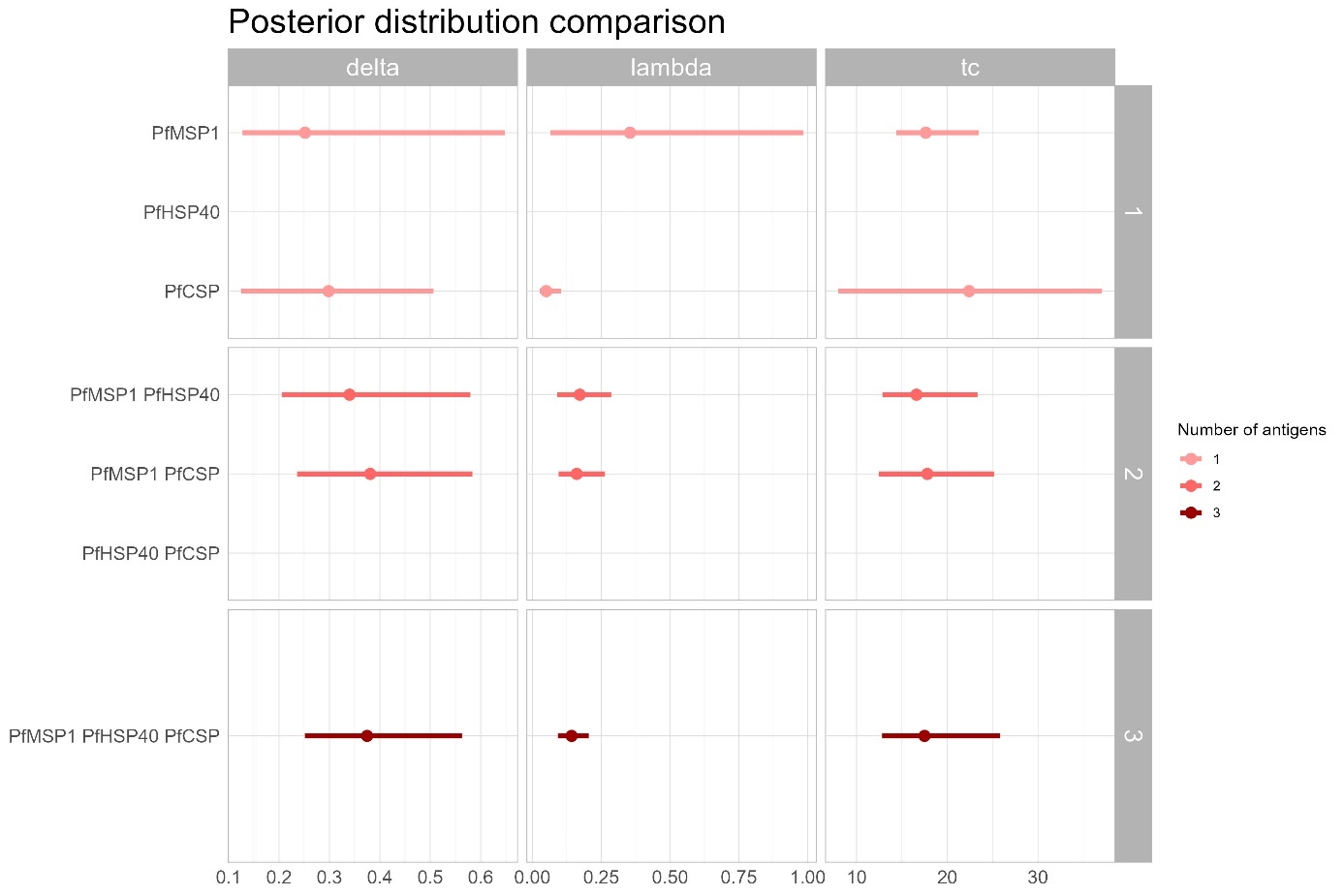

***Figure S18: Posterior distribution comparison.*** *Posterior distribution of the three parameters that have been compared to validation data: sero-incidence, time and magnitude of sharp drop in transmission. Model presented in this plot are all possible combinations of PfMSP1, PfHSP40 and PfCSP. Only models that converged are presented.*

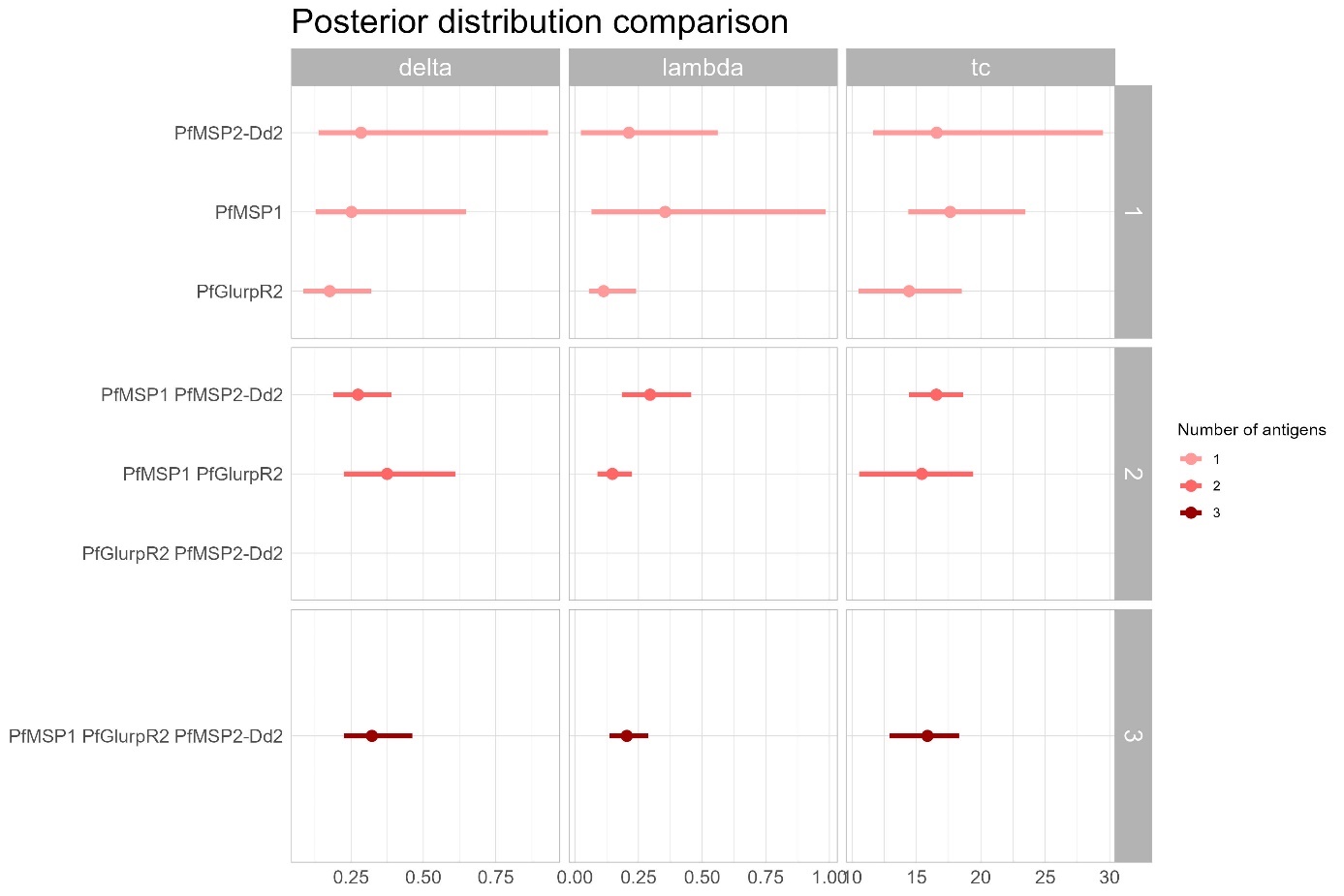

***Figure S19: Posterior distribution comparison.*** *Posterior distribution of the three parameters that have been compared to validation data: sero-incidence, time and magnitude of sharp drop in transmission. Model presented in this plot are all possible combinations of PfMSP1, PfGlurpR2 and PfMSP2-Dd2. Only models that converged are presented.*

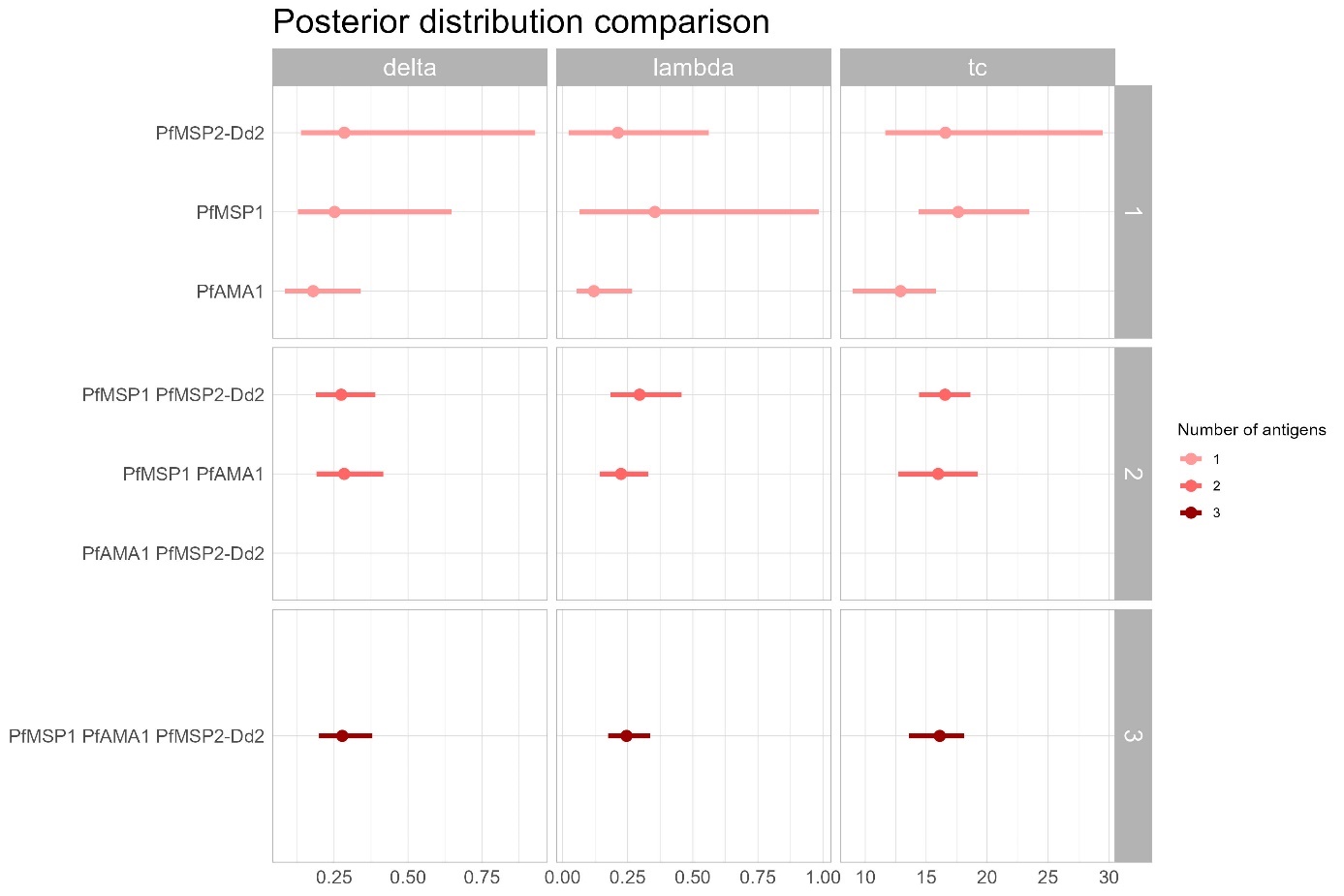

***Figure S20: Posterior distribution comparison.*** *Posterior distribution of the three parameters that have been compared to validation data: sero-incidence, time and magnitude of sharp drop in transmission. Model presented in this plot are all possible combinations of PfMSP1, PfAMA1 and PfMSP2-Dd2. Only models that converged are presented.*

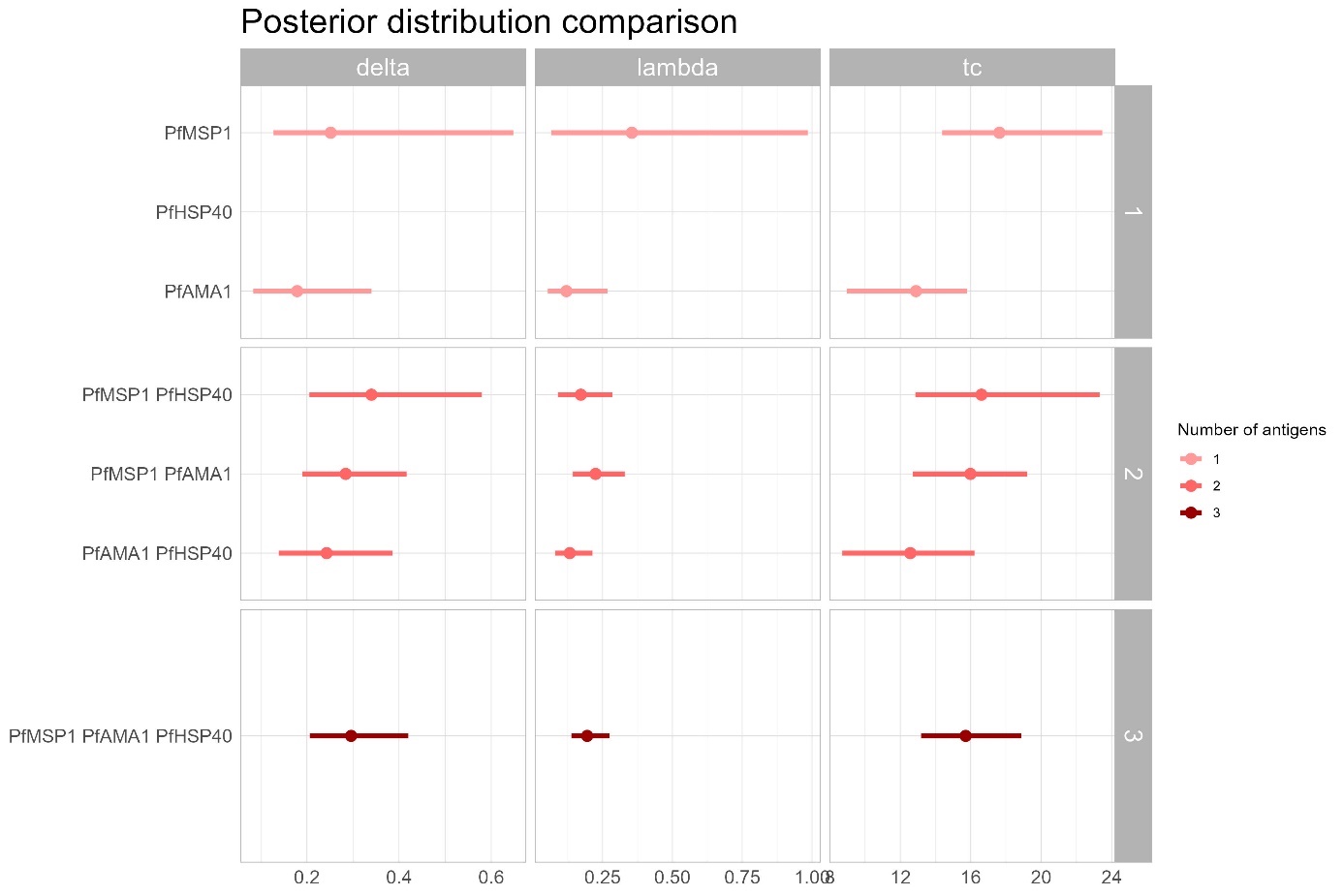

***Figure S21: Posterior distribution comparison.*** *Posterior distribution of the three parameters that have been compared to validation data: sero-incidence, time and magnitude of sharp drop in transmission. Model presented in this plot are all possible combinations of PfMSP1, PfAMA1 and PfHSP40. Only models that converged are presented.*

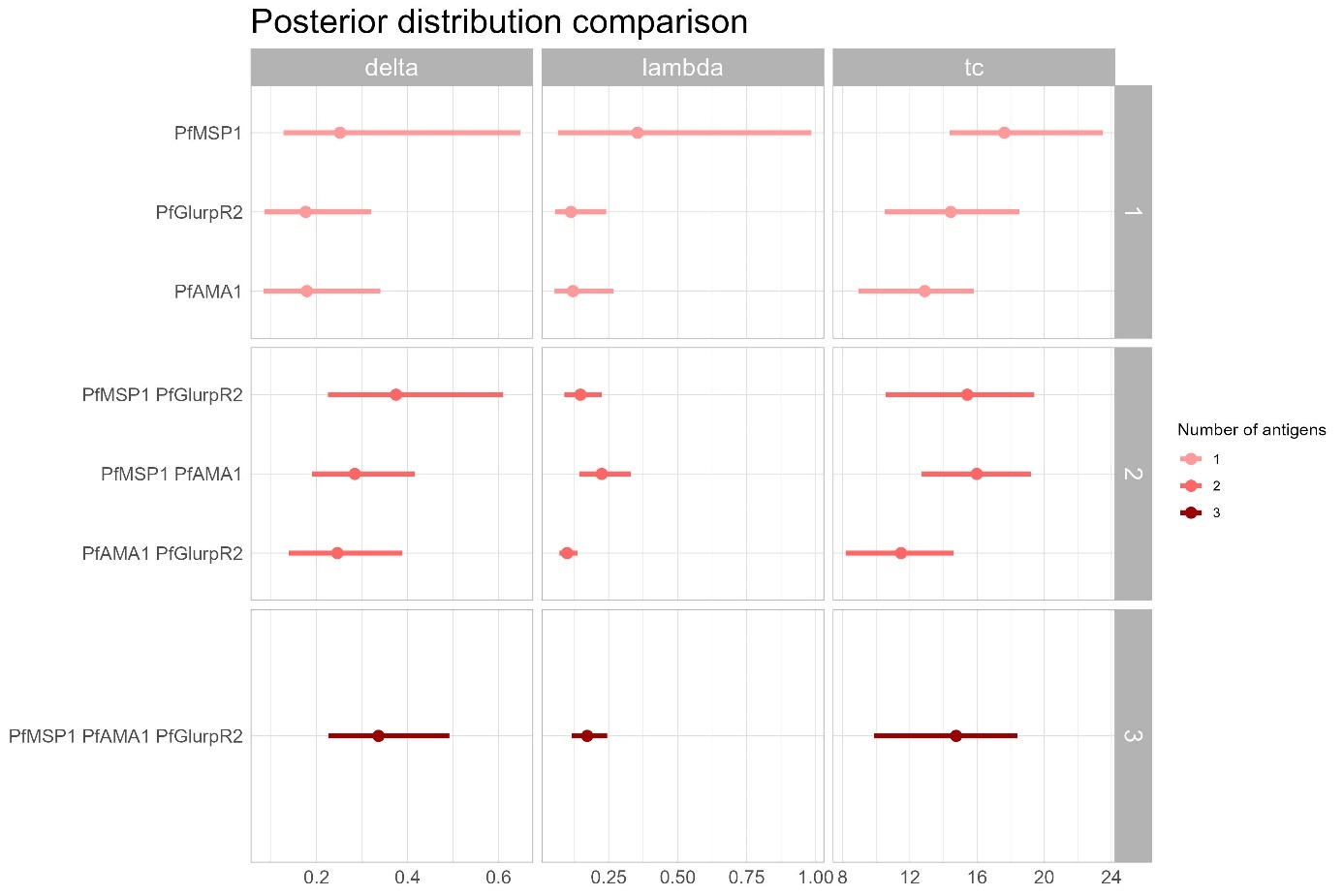

***Figure S22: Posterior distribution comparison.*** *Posterior distribution of the three parameters that have been compared to validation data: sero-incidence, time and magnitude of sharp drop in transmission. Model presented in this plot are all possible combinations of PfMSP1, PfAMA1 and PfGlurpR2. Only models that converged are presented.*

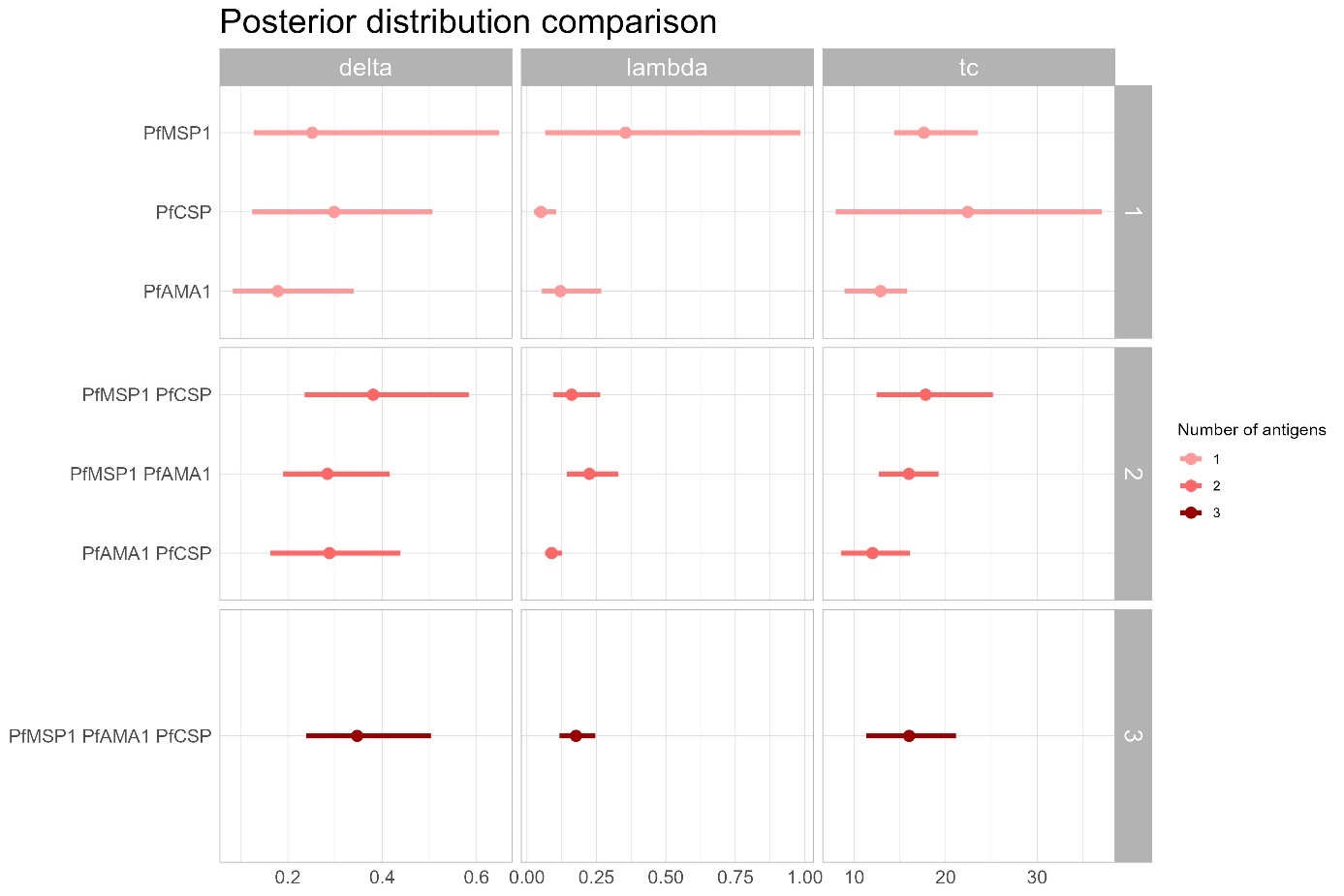

***Figure S23: Posterior distribution comparison.*** *Posterior distribution of the three parameters that have been compared to validation data: sero-incidence, time and magnitude of sharp drop in transmission. Model presented in this plot are all possible combinations of PfMSP1, PfAMA1 and PfCSP. Only models that converged are presented.*
